# Latent Class Analysis Identifies Pulmonary Function Trajectory Phenotypes in Lung Transplant Recipients with Chronic Allograft Dysfunction

**DOI:** 10.64898/2026.04.22.26351501

**Authors:** Megan L. Neely, Daniel M. Wojdyla, Hwanhee Hong, Peijin Wang, Michaela R. Anderson, Katelyn Arroyo, John A. Belperio, Luke Benvenuto, Marie Budev, Michael P. Combs, Gundeep Dhillon, Jesse Y. Hsu, Laurel Kalman, Tereza Martinu, John McDyer, Michelle Oyster, Krishna Pandya, John M. Reynolds, Jeeyon G. Rim, David W. Roe, Pali D. Shah, Jonathan P. Singer, Lianne G. Singer, Laurie D. Snyder, Wayne Tsuang, S. Samuel Weigt, Jason D. Christie, Scott M. Palmer, Jamie L. Todd

## Abstract

**Background:** We aimed to identify data-driven FEV1 trajectory phenotypes post-chronic lung allograft dysfunction (CLAD), relate these phenotypes to patient factors and future graft loss, and develop a classification approach for prospective patients.

**Methods:** We studied adult first lung recipients with probable CLAD from two prospective multicenter cohorts: CTOT-20 (n=206) and LTOG (n=1418). FEV1 trajectories over the first nine months post-CLAD were characterized using joint latent class mixed models, jointly modelling time-to-graft loss to account for informative censoring. Models were fit independently in both cohorts and also only among LTOG bilateral recipients. A classification and regression tree (CART) model was derived in LTOG bilateral recipients and applied to CTOT-20 bilateral recipients.

**Findings:** Four distinct early FEV1 trajectory classes were identified in CTOT-20, with large differences in nine-month graft loss (72·3%, 31·1%, 2·2%, 0%). In LTOG, similar trajectory patterns were reproduced, with an additional class demonstrating early post-CLAD FEV1 improvement. Among bilateral recipients, trajectory classes showed a clear risk gradient, including a high-risk class with 100% graft loss and a low-risk class with no early graft loss. A CART model incorporating clinical and spirometric variables demonstrated good discrimination in LTOG bilateral recipients (multiclass AUC 0·85) and consistent class assignment and trajectory patterns when applied to CTOT-20.

**Interpretation:** We identified reproducible, clinically meaningful early post-CLAD FEV1 trajectory phenotypes with differential graft loss risk. These phenotypes and a pragmatic classification tool may support risk stratification, trial enrichment, and improved prognostication for patients and clinicians.

**Funding:** National Institutes of Health, Cystic Fibrosis Foundation

## INTRODUCTION

Lung transplantation is a life-extending therapy for patients with end-stage lung disease; however, long-term survival remains limited to a median of about seven years.^1^ Chronic lung allograft dysfunction (CLAD) is the leading cause of late posttransplant mortality and represents a major barrier to improved lung transplant success,^1,2^ yet there are no approved therapies. CLAD is characterized by immune-mediated fibrosis of the small airways and/or lung parenchyma and is clinically diagnosed by a sustained decline in pulmonary function.^2^ In 2019, an International Society for Heart and Lung Transplantation (ISHLT) consensus updated the CLAD diagnostic approach to incorporate CLAD certainty categories (i.e., probable, definite) based on duration of lung function decline.^2^ Probable CLAD was proposed as an entry point for clinical trials to enable treatment earlier in the disease course; however, the natural history of lung function progression after probable CLAD has not been well defined, resulting in uncertainty about how to best define inclusion or stratification criteria to create homogenous populations for interventional studies.

In clinical practice, a wide spectrum of lung function trajectories after CLAD is observed. Studies describing lung function progression post-CLAD are few and limited to single-center retrospective cohorts that predate the updated CLAD consensus definitions.^3–5^ Recent efforts to define clinical CLAD phenotypes have provided some prognostic insight.^6–9^ However, these recognized phenotypes do not fully account for the wide variability in disease trajectories observed in practice. These limitations along with the critical need to accelerate therapeutics for CLAD underscore the need for a novel, data-driven strategy to identify clinically meaningful lung function trajectory subgroups within CLAD that can inform trial design and improve patient prognostication.

Latent class analysis (LCA) of longitudinal post-CLAD pulmonary function data provides a promising approach to address this unmet need, revealing unobserved subgroups that contribute to heterogenous progression outcomes within lung recipients with CLAD.^10,11^ We leveraged two multicenter lung transplant cohort studies, the Clinical Trials in Organ Transplantation (CTOT-20) study and the Lung Transplant Outcomes Group (LTOG) study, to identify latent classes of CLAD progression using serial FEV1 measurements following CLAD onset. We additionally assessed the association between these classes and future graft loss and applied a machine learning approach to generate a clinically interpretable decision rule that could be used at CLAD onset or shortly thereafter to inform the early post-CLAD progression course a patient may experience.

## METHODS

### Study population and CLAD ascertainment

The population was drawn from two prospective observational lung recipient cohort studies, CTOT-20 and LTOG. CTOT-20 was designed to identify clinical and biological risk factors for CLAD and enrolled 803 newly transplanted first adult lung recipients at five North American centers between 2015–2018.^12^ Patients underwent prospective identification of probable CLAD over a median follow-up of 3·4 (2·0, 4·3) years, including center-level adjudication to confirm the absence of confounding conditions.^13^ Probable CLAD was defined as a ≥20% decline in the FEV1 as compared with the average of the two best posttransplant FEV1s measured at least three weeks apart (posttransplant baseline) that was sustained for a minimum of 21 days in the absence of clinical confounders.^2^ The probable CLAD onset date was defined as the date of the first threshold FEV1 decline. Lung recipients in CTOT-20 who developed probable CLAD by these pulmonary function test (PFT) criteria were included in the current analysis (n=206).^13^

LTOG is a prospective observational cohort study designed to investigate early and late posttransplant complications.^14,15^ LTOG enrolled 2448 newly transplanted lung recipients at 10 US centers between 2007–2018. Multi-organ or lobar recipients (n=34) were excluded as were patients with incomplete PFT data (n=28, representing all patients from a single center). The remaining 2386 patients were followed until death, retransplantation, early termination or June 30, 2023, for a median follow-up of 6·1 (3·4, 8·1) years. PFT measures performed as part of clinical care were entered into a central database and the first occurrence of probable CLAD was identified as the first ≥20% decline in the FEV1 as compared with the posttransplant baseline that was sustained for a minimum of 21 days. As for CTOT-20, the date of CLAD onset was defined as the date of the first threshold FEV1 decline. Lung recipients in LTOG who developed probable CLAD by these PFT criteria were included in the current analysis (n=1418).^15^ One center contributed patients to both CTOT-20 and LTOG with the overlap representing only 2·6% (n=37/1418) of the LTOG CLAD cohort. All patients received clinical management according to center practices. The study was approved by the Duke Institutional Review Board (Pro00113359).

### Features of interest at CLAD onset

Clinical features of interest at CLAD onset included CLAD stage, timing of CLAD onset, and several physiological measures. CLAD stage (one to four) was defined according to ISHLT guidelines, based on the degree of FEV1 decline present on the CLAD-onset PFT compared with the posttransplant baseline FEV1.^2^ Early-onset CLAD was defined as CLAD occurring within two years of transplantation.^3^ Decline in forced vital capacity (FVC) at CLAD onset (i.e., FVC loss), which may identify patients with a restrictive ventilatory defect, was defined as CLAD onset FVC/FVC_Baseline_ <0·8 where FVC_Baseline_ was the average of the two FVC measurements that paired with the two best posttransplant FEV1s used in the CLAD calculation.^8,9^ Lastly, we examined the FEV1/FVC ratio from the CLAD onset PFT; specifically, FEV1/FVC <0·7 as this is the threshold used to define obstructive physiology.

### Statistical analysis

Demographics, transplant characteristics, and clinical features at CLAD onset were summarized using median (first, third quartile [Q1, Q3]) for continuous variables and counts with percentages for categorical variables. To uncover distinct classes of patients with similar lung function trajectories following CLAD onset we employed LCA, a mixture modeling technique used to identify unobserved subpopulations based on longitudinal patterns. LCA has been successfully applied in lung transplant research including the identification of phenotypes in primary graft dysfunction (PGD),^16^ and in studies characterizing FEV1 trajectories beginning at transplantation,^17^ as well as in non-transplant lung diseases.^18^ The analysis included serial FEV1 measures within the first nine months following CLAD diagnosis. This time window was selected because it represented the period with the densest longitudinal PFT data. Beyond nine months, increasing rates of graft loss (defined as death or retransplantation) resulted in a higher proportion of censored FEV1 data, limiting the reliability of trajectory estimation and introducing potential non-random censoring. To account for this, a joint modeling framework was implemented, enabling simultaneous estimation of the longitudinal FEV1 trajectory and time to graft loss.^10,11^

The joint LCA model included two components: 1) a longitudinal submodel characterizing FEV1 trajectories using time since CLAD onset as a covariate with subject-specific random intercepts and slopes and a restricted cubic spline on time to allow for non-linear trends, and 2) a survival submodel for time to graft loss. The primary component of interest in this analysis was the longitudinal submodel; the survival submodel was included to account for non-random censoring of the longitudinal process that might otherwise have resulted in bias to the estimates of the longitudinal trajectory parameters. Covariates included in the survival component were those that may be plausibly related to post-CLAD graft loss including age at CLAD onset, CLAD timing (early vs. not), and transplant type (bilateral vs. single). The model was estimated using the *lcmm* package in R.^19,20^ Prior to model fitting, preliminary analyses were conducted to determine the appropriate correlation structure for the repeated FEV1 measures, the functional form of time in the longitudinal submodel, the shape of the hazard function in the survival component, and potential differences in hazard functions across latent classes (**Supplementary Material**).

Model selection was guided by the Bayesian Information Criterion (BIC) and the clinical interpretability of the derived classes. Posterior class membership probabilities were used to assess classification certainty, with mean posterior probabilities >0·80 indicating stable class assignment. Class size was also considered in model selection, with a general threshold of no fewer than 5–10% of the cohort assigned to any class. Each latent class was described graphically using estimated mean FEV1 trajectories and qualitatively using baseline characteristics including demographics, native lung disease, transplant type, and clinical features of interest at CLAD onset. Between group comparisons were performed using the Kruskal-Wallis test for continuous variables and chi-square or Fisher’s exact test for categorical variables. To evaluate graft loss across the identified latent classes, we used Kaplan-Meier analyses to estimate the cumulative incidence of post-CLAD graft loss, stratified by CLAD latent class. All modeling was performed in the CTOT-20 and LTOG datasets independently. Additionally, the LCA modeling was repeated considering only bilateral lung recipients in LTOG as this is a clinically relevant subgroup.

To identify predictors of latent class membership and generate a clinically interpretable decision rule for prospective recipients with CLAD, we applied Classification and Regression Tree (CART) modeling in the LTOG bilateral lung recipient CLAD population using the *<u>rpart</u>* package in R.^21^ CART is a nonparametric machine learning approach that recursively partitions the data to create homogeneous outcome groups based on predictor variables.^22^ We used the assigned latent class membership as the dependent variable and included candidate demographic, clinical, and spirometric predictors in the model. Two models were considered: one with select sociodemographic and clinical factors known at the time of CLAD diagnosis including timing of CLAD onset and CLAD stage (clinical only) and one with these factors along with select spirometric variables (clinical + spirometry) (**Table E1**). Tree construction employed the Gini impurity criterion to evaluate node splits. To reduce overfitting, we used 10-fold cross-validation, selecting the tree with the lowest cross-validated error within one standard error of the minimum. Variable importance measures were extracted to identify the most influential predictors. Model accuracy was evaluated in the derivation cohort using cross-validated overall classification accuracy, class-specific sensitivity and specificity, and the multiclass area under the receiver operating characteristic curve (AUC).

The CART model derived in the LTOG bilateral cohort was then applied to the CTOT-20 bilateral cohort to understand how well it performed in this new population. The performance of the model in the CTOT-20 bilateral recipients was evaluated by 1) assessing the distribution of CTOT-20 bilateral recipients predicted to be in each of the four classes identified in LTOG bilateral recipients, and 2) assessing if the estimated post-CLAD FEV1 trajectory of the CTOT-20 bilateral recipients in each predicted class was similar to the trajectories from each class identified in LTOG bilateral recipient CLAD population. All tests were two-sided with a significance threshold of α=0·05. All analyses were completed in SAS version 9·4 (SAS Institute, Inc., Cary, NC) or R version 4·4·2.^23^

## RESULTS

### Cohort characteristics

**Table 1** shows the clinical characteristics of the CTOT-20 and LTOG CLAD cohorts. Recipient demographics at transplant, transplant type distribution, grade three PGD rates, and posttransplant baseline FVC and FEV1 values were similar in both groups. The median number of PFTs available within the nine months following CLAD onset was also similar (5 [4, 7] for CTOT-20 and 5 [3, 7] for LTOG). Considering CLAD onset features, patients with CLAD in CTOT-20 were numerically more likely to have early onset CLAD and CLAD of stage ≥2 at onset. FVC loss was present in about one-third of patients in both cohorts. Death or retransplant occurred within nine months of CLAD onset in 19·9% of the CTOT-20 CLAD cohort and 12·4% of the LTOG CLAD cohort.

**Table 1.**
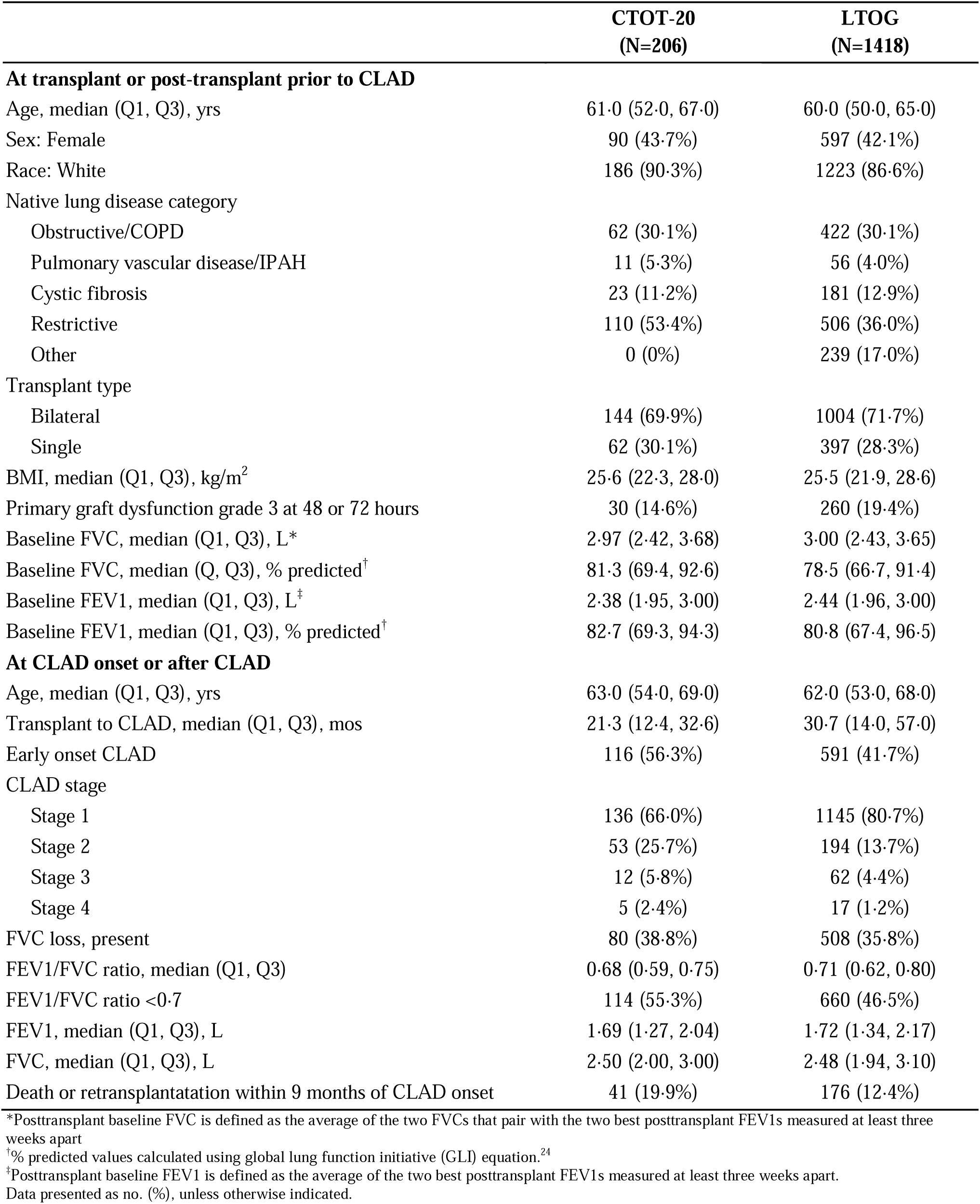
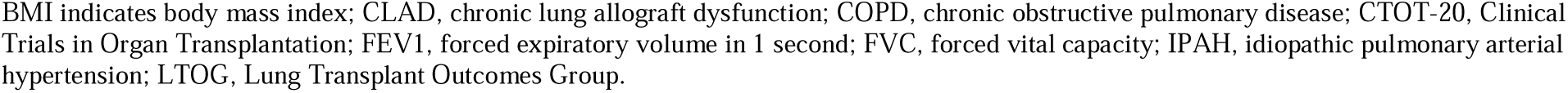
Characteristics of the CTOT-20 and LTOG probable CLAD cohorts.

|  | **CTOT-20**<br>**(N=206)** | **LTOG**<br>**(N=1418)** |
| --- | --- | --- |
| **At transplant or post-transplant prior to CLAD** |  |  |
| Age, median (Q1, Q3), yrs | 61.0 (52.0, 67.0) | 60.0 (50.0, 65.0) |
| Sex: Female | 90 (43.7%) | 597 (42.1%) |
| Race: White | 186 (90.3%) | 1223 (86.6%) |
| Native lung disease category |  |  |
| Obstructive/COPD | 62 (30.1%) | 422 (30.1%) |
| Pulmonary vascular disease/IPAH | 11 (5.3%) | 56 (4.0%) |
| Cystic fibrosis | 23 (11.2%) | 181 (12.9%) |
| Restrictive | 110 (53.4%) | 506 (36.0%) |
| Other | 0 (0%) | 239 (17.0%) |
| Transplant type |  |  |
| Bilateral | 144 (69.9%) | 1004 (71.7%) |
| Single | 62 (30.1%) | 397 (28.3%) |
| BMI, median (Q1, Q3), kg/m<sup>2</sup> | 25.6 (22.3, 28.0) | 25.5 (21.9, 28.6) |
| Primary graft dysfunction grade 3 at 48 or 72 hours | 30 (14.6%) | 260 (19.4%) |
| Baseline FVC, median (Q1, Q3), L* | 2.97 (2.42, 3.68) | 3.00 (2.43, 3.65) |
| Baseline FVC, median (Q, Q3), % predicted<sup>†</sup> | 81.3 (69.4, 92.6) | 78.5 (66.7, 91.4) |
| Baseline FEV1, median (Q1, Q3), L<sup>‡</sup> | 2.38 (1.95, 3.00) | 2.44 (1.96, 3.00) |
| Baseline FEV1, median (Q1, Q3), % predicted<sup>†</sup> | 82.7 (69.3, 94.3) | 80.8 (67.4, 96.5) |
| **At CLAD onset or after CLAD** |  |  |
| Age, median (Q1, Q3), yrs | 63.0 (54.0, 69.0) | 62.0 (53.0, 68.0) |
| Transplant to CLAD, median (Q1, Q3), mos | 21.3 (12.4, 32.6) | 30.7 (14.0, 57.0) |
| Early onset CLAD | 116 (56.3%) | 591 (41.7%) |
| CLAD stage |  |  |
| Stage 1 | 136 (66.0%) | 1145 (80.7%) |
| Stage 2 | 53 (25.7%) | 194 (13.7%) |
| Stage 3 | 12 (5.8%) | 62 (4.4%) |
| Stage 4 | 5 (2.4%) | 17 (1.2%) |
| FVC loss, present | 80 (38.8%) | 508 (35.8%) |
| FEV1/FVC ratio, median (Q1, Q3) | 0.68 (0.59, 0.75) | 0.71 (0.62, 0.80) |
| FEV1/FVC ratio <0.7 | 114 (55.3%) | 660 (46.5%) |
| FEV1, median (Q1, Q3), L | 1.69 (1.27, 2.04) | 1.72 (1.34, 2.17) |
| FVC, median (Q1, Q3), L | 2.50 (2.00, 3.00) | 2.48 (1.94, 3.10) |
| Death or retransplantation within 9 months of CLAD onset | 41 (19.9%) | 176 (12.4%) |

\*Posttransplant baseline FVC is defined as the average of the two FVCs that pair with the two best posttransplant FEV1s measured at least three weeks apart
<sup>†</sup>% predicted values calculated using global lung function initiative (GLI) equation.<sup>24</sup>
<sup>‡</sup>Posttransplant baseline FEV1 is defined as the average of the two best posttransplant FEV1s measured at least three weeks apart.
Data presented as no. (%), unless otherwise indicated.
BMI indicates body mass index; CLAD, chronic lung allograft dysfunction; COPD, chronic obstructive pulmonary disease; CTOT-20, Clinical Trials in Organ Transplantation; FEV1, forced expiratory volume in 1 second; FVC, forced vital capacity; IPAH, idiopathic pulmonary arterial hypertension; LTOG, Lung Transplant Outcomes Group.

### Identification of early post-CLAD FEV1 trajectories in CTOT-20

To identify distinct early post-CLAD FEV1 trajectories in CTOT-20 patients with prospectively adjudicated probable CLAD, latent class models with two to four classes were evaluated. Model fit statistics are shown in **Table E2**. While the two-class model had the lowest BIC, both the three- and four-class solutions identified clinically meaningful classes of FEV1 trajectories and maintained mean posterior class membership probabilities >0·8 (**Figures E1 and E2, Table 2**). The four-class solution was selected for further analysis based on favorable fit, entropy, and posterior probability metrics, as well as the ability to capture a broader range of clinically relevant disease patterns. The estimated parameters for the four class solution are specified in **Table E3**.

**Table 2.** Clinical characteristics and CLAD onset features by class membership in the CTOT-20 probable CLAD cohort.

|  | **Class 1**<br>**(N=47)** | **Class 2**<br>**(N=16)** | **Class 3**<br>**(N=90)** | **Class 4**<br>**(N=53)** | **p-value** |
| --- | --- | --- | --- | --- | --- |
| **At transplant or posttransplant prior to CLAD** |  |  |  |  |  |
| Age at transplant, median (Q1, Q3), yrs | 55.0 (35.0, 65.0) | 61.0 (54.0, 64.5) | 62.0 (57.0, 68.0) | 60.0 (51.0, 64.0) | 0.005 |
| Sex: Female | 23 (48.9%) | 14 (87.5%) | 37 (41.1%) | 16 (30.2%) | <001 |
| Race: White | 40 (85.1%) | 12 (75.0%) | 84 (93.3%) | 50 (94.3%) | 0.055 |
| Native lung disease category |  |  |  |  | 0.003 |
| Obstructive/COPD | 15 (31.9%) | 6 (37.5%) | 23 (25.6%) | 18 (34.0%) |  |
| Pulmonary vascular disease/IPAH | 4 (8.5%) | 2 (12.5%) | 1 (1.1%) | 4 (7.5%) |  |
| Cystic fibrosis | 11 (23.4%) | 1 (6.3%) | 4 (4.4%) | 7 (13.2%) |  |
| Restrictive | 17 (36.2%) | 7 (43.8%) | 62 (68.9%) | 24 (45.3%) |  |
| Transplant type |  |  |  |  | <001 |
| Bilateral | 36 (76.6%) | 11 (68.8%) | 48 (53.3%) | 49 (92.5%) |  |
| Single | 11 (23.4%) | 5 (31.3%) | 42 (46.7%) | 4 (7.5%) |  |
| BMI, median (Q1, Q3), kg/m<sup>2</sup> | 24.6 (20.0, 27.2) | 25.3 (23.4, 26.2) | 26.5 (23.2, 29.1) | 25.4 (22.2, 27.8) | 0.035 |
| Primary graft dysfunction grade 3 at 48 or 72 hours | 9 (19.1%) | 4 (25.0%) | 13 (14.4%) | 4 (7.5%) | 0.232 |
| Baseline FVC, median (Q1, Q3), L* | 2.95 (2.45, 3.59) | 2.12 (1.70, 2.42) | 2.76 (2.40, 3.17) | 3.79 (3.13, 4.40) | <001 |
| Baseline FVC, median (Q1, Q3), % predicted<sup>†</sup> | 82.0 (70.7, 91.9) | 68.0 (50.2, 86.2) | 78.4 (66.2, 89.0) | 90.0 (77.2, 98.3) | <001 |
| Baseline FEV1, median (Q1, Q3), L<sup>‡</sup> | 2.48 (2.04, 3.13) | 1.50 (1.23, 1.91) | 2.24 (1.89, 2.50) | 3.15 (2.66, 3.64) | <001 |
| Baseline FEV1, median (Q1, Q3), % predicted<sup>‡</sup> | 84.2 (69.0, 91.8) | 64.0 (48.8, 80.8) | 79.2 (66.8, 88.1) | 93.4 (82.7, 105.8) | <001 |
| **At CLAD onset or after CLAD** |  |  |  |  |  |
| Age at CLAD onset, median (Q1, Q3), yrs | 58.0 (36.0, 67.0) | 62.5 (55.5, 66.5) | 64.0 (60.0, 71.0) | 61.0 (53.0, 66.0) | 0.003 |
| Transplant to CLAD, median (Q1, Q3), mos | 20.2 (14.7, 31.2) | 16.4 (11.4, 24.9) | 23.3 (12.4, 35.3) | 22.2 (9.9, 32.5) | 0.533 |
| Early onset CLAD | 31 (66.0%) | 11 (68.8%) | 46 (51.1%) | 28 (52.8%) | 0.258 |
| CLAD stage at onset |  |  |  |  | <001 |
| Stage 1 | 28 (59.6%) | 3 (18.8%) | 63 (70.0%) | 42 (79.2%) |  |
| Stage 2 | 15 (31.9%) | 5 (31.3%) | 23 (25.6%) | 10 (18.9%) |  |
| Stage 3 | 3 (6.4%) | 4 (25.0%) | 4 (4.4%) | 1 (1.9%) |  |
| Stage 4 | 1 (2.1%) | 4 (25.0%) | 0 (0.0%) | 0 (0.0%) |  |
| FVC loss at CLAD onset | 21 (44.7%) | 10 (62.5%) | 32 (35.6%) | 17 (32.1%) | 0.118 |
| FEV1/FVC ratio at CLAD onset, median (Q1, Q3) | 0.68 (0.58, 0.76) | 0.50 (0.44, 0.67) | 0.67 (0.59, 0.74) | 0.73 (0.64, 0.79) | 0.001 |
| FEV1/FVC ratio at CLAD onset <0.7 | 26 (55.3%) | 13 (81.3%) | 52 (57.8%) | 23 (43.4%) | 0.055 |
| FEV1 at CLAD onset, median (Q1, Q3), L | 1.69 (1.17, 2.04) | 0.71 (0.60, 0.90) | 1.59 (1.33, 1.83) | 2.35 (1.97, 2.63) | <001 |
| FVC at CLAD onset, median (Q1, Q3), L | 2.47 (2.01, 2.93) | 1.40 (1.11, 1.95) | 2.28 (1.94, 2.73) | 3.26 (2.70, 3.76) | <001 |
| Death or retransplantation within 9 months | 34 (72.3%) | 5 (31.3%) | 2 (2.2%) | 0 (0.0%) | <001 |
| Mean of posterior probabilities | 0.86 | 0.81 | 0.83 | 0.87 |  |

\*Posttransplant baseline FVC is defined as the average of the two FVCs that pair with the two best posttransplant FEV1s measured at least three weeks apart
<sup>†</sup>% predicted values calculated using global lung function initiative (GLI) equation.<sup>24</sup>
<sup>‡</sup>Posttransplant baseline FEV1 is defined as the average of the two best posttransplant FEV1s measured at least three weeks apart.
Data presented as no. (%), unless otherwise indicated.
BMI indicates body mass index; CLAD, chronic lung allograft dysfunction; COPD, chronic obstructive pulmonary disease; FEV1, forced expiratory volume in 1 second; FVC, forced vital capacity; IPAH, idiopathic pulmonary arterial hypertension.

The mean predicted post-CLAD FEV1 trajectory for each of the four classes is shown in **Figure 1A** while individual FEV1 trajectories using observed data are illustrated in **Figure E3**. Many clinical characteristics differed significantly across the classes (**Table 2**) as did the cumulative incidence of graft loss (**Figure 1B**). Patients in class one (n=47/206 [22·8%]), experienced the greatest FEV1 decline and had the highest rates of graft loss (72·3%) in the nine months following CLAD onset (**Table 2**, **Figure 1B**). These patients with the worst prognosis were younger, more likely to have cystic fibrosis native lung disease, and more often had CLAD stage ≥2 at onset. Patients in class two (n=16/206 [7·8%]) demonstrated intermediate prognosis. Nearly one-third (31·1%) of patients in class two experienced graft loss within nine months after CLAD. Patients in class two were predominantly female (87·5%) with subnormal baseline lung function (median baseline FVC and FEV1 of 68% and 64% predicted) and marked lung function impairment at CLAD onset (**Table 2**).

**Figure 1.**
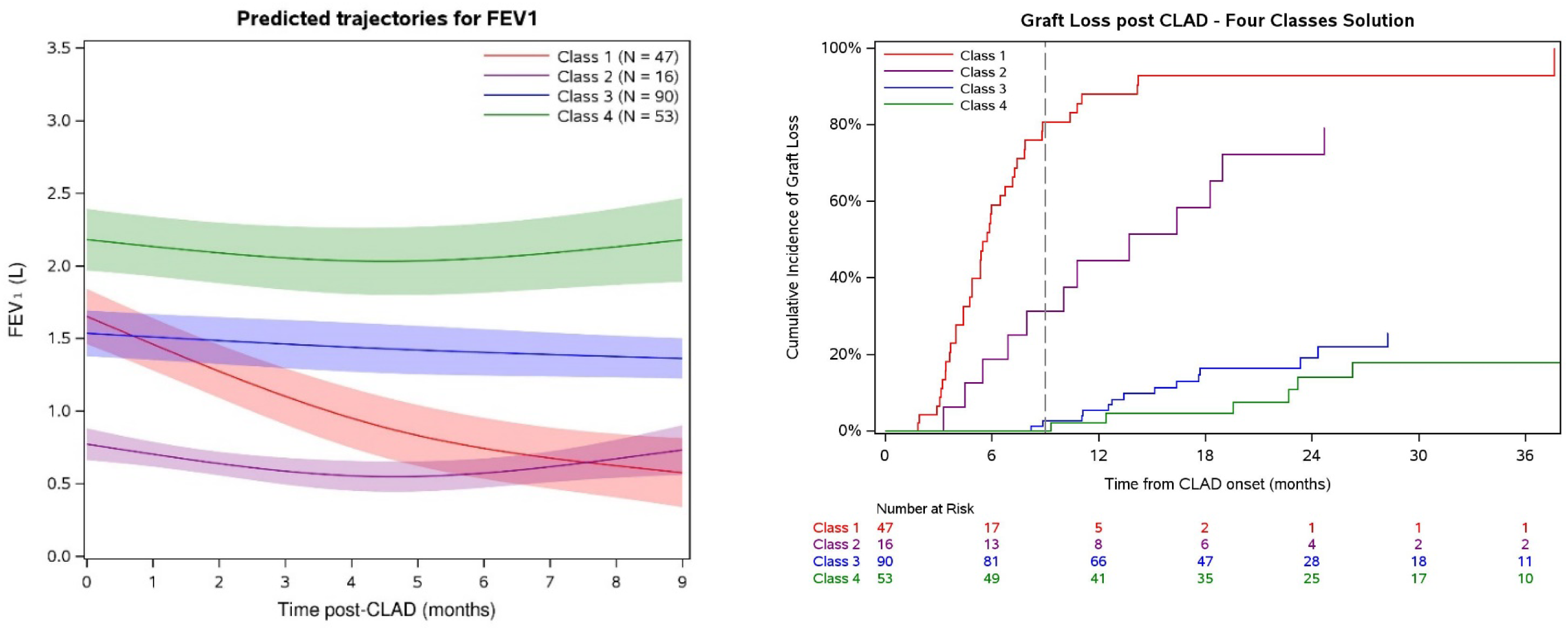
In the CTOT-20 CLAD cohort, **A)** estimated mean FEV1 trajectory in the nine months following CLAD onset for each of the four latent classes and **B)** cumulative incidence of graft loss after CLAD by class membership. Dashed line represents nine months post-CLAD.

In contrast, patients in classes three (n=90/206 [43·7%]) and four (n=53/206 [25·7%]) had better prognosis (**Table 2**, **Figure 1B**). Only 2·2% of patients in class three and no patients in class four had graft loss within nine months of CLAD. Patients in class three were more likely to have restrictive native lung disease, be single lung transplant recipients, and have less stage one CLAD at onset. Patients in class four were more often male and bilateral lung recipients with stage one CLAD at onset. These patients had the highest FEV1 values both at posttransplant baseline and at CLAD diagnosis and had relatively stable lung function in the early post-CLAD period.

#### Identification of early post-CLAD FEV1 trajectories in LTOG

To determine whether similar FEV1 trajectory patterns occur after probable CLAD in a largely independent cohort, the same modeling approach was applied to the LTOG CLAD cohort. The LTOG cohort is larger than the CTOT-20 cohort and includes longer follow-up periods, which may provide a broader representation of CLAD disease heterogeneity. Models with two to six classes were considered. Fit statistics suggested a five-class solution as best (**Table E4**) and estimated parameters of the five-class solution are given in **Table E5**. Classes one to four showed early post-CLAD FEV1 trajectory patterns that were consistent with those observed in CTOT-20 (**Figure 2A, Figure E4**). As in CTOT-20, patients in classes one (n=129/1418 [9·1%]) and two (n=220/1418 [15·5%]) experienced a higher cumulative incidence of graft loss compared with those in classes three (n=603/2428 [42·5%]) and four (n=101/1418 [7·1%]) (**Figure 2B**).

**Figure 2.**
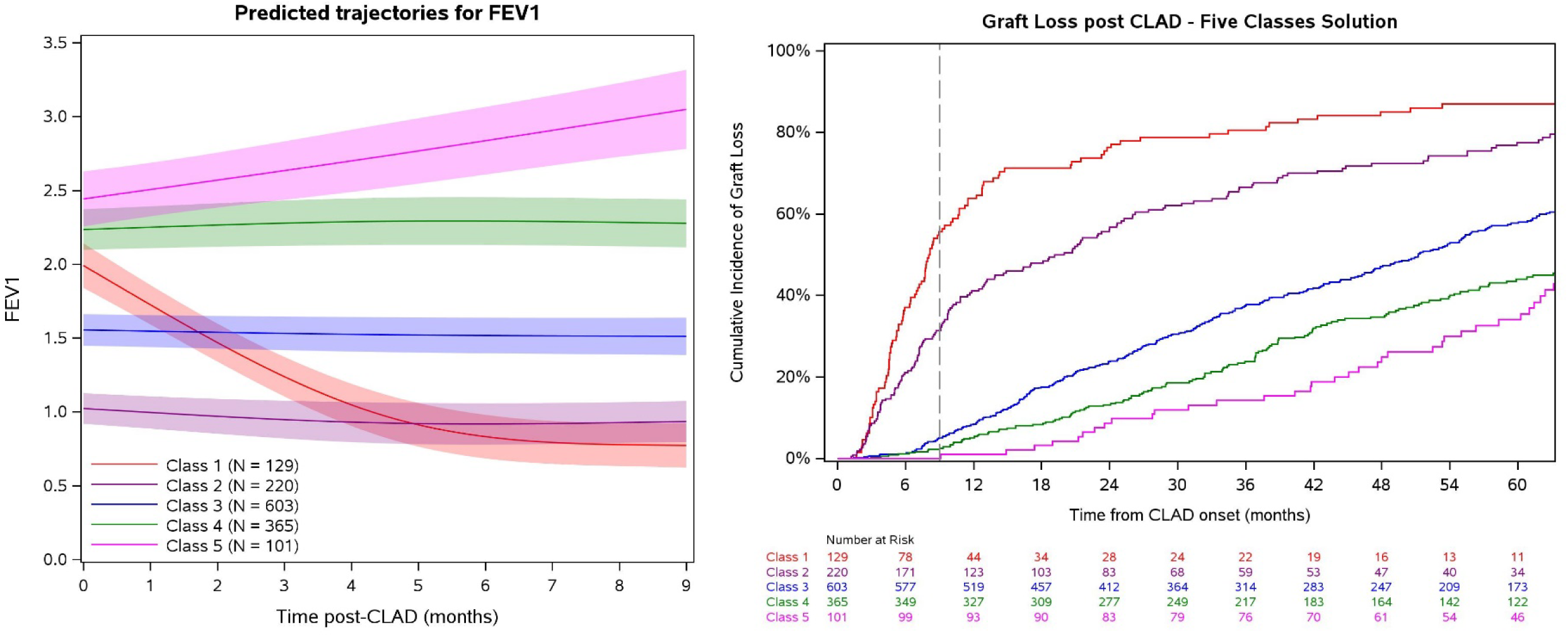
In the LTOG CLAD cohort, **A)** estimated mean FEV1 trajectory in the nine months following CLAD onset for each of the five latent classes and **B)** cumulative incidence of graft loss after CLAD by class membership. Dashed line represents nine months post-CLAD.

The clinical characteristics associated with each class are summarized in **Table 3**. The pattern of clinical characteristics observed across classes one to four were generally consistent with those found in CTOT-20. Patients in class one tended to be younger, more frequently had cystic fibrosis, and exhibited a shift toward stage two or three CLAD at onset. Class two was characterized by a predominance of females with poor baseline lung function and severe onset CLAD. These patients also had higher rates of severe PGD. Individuals in class three were more likely to have restrictive native disease, be single lung recipients, and have stage one CLAD. Class four comprised predominantly males, bilateral lung recipients, individuals with good baseline lung function, and stage one CLAD at onset. In the larger LTOG cohort, an additional group (class five) was identified in which the mean FEV1 trajectory demonstrated improvement. Nearly all these patients were male and bilateral lung recipients with median baseline lung function values at or approaching 100% predicted.

**Table 3.** Clinical characteristics and CLAD onset features by class membership in the LTOG probable CLAD cohort.

|  | **Class 1**<br>**(N=129)** | **Class 2**<br>**(N=220)** | **Class 3**<br>**(N=603)** | **Class 4**<br>**(N=365)** | **Class 5**<br>**(N=101)** | **p-value** |
| --- | --- | --- | --- | --- | --- | --- |
| **At transplant or posttransplant prior to CLAD** |  |  |  |  |  |  |
| Age at transplant, median (Q1, Q3), yrs | 55.0 (32.0, 61.0) | 60.0 (51.0, 65.5) | 61.0 (54.0, 66.0) | 59.0 (48.0, 65.0) | 55.0 (40.0, 62.0) | <.001 |
| Sex: Female | 50 (38.8%) | 163 (74.4%) | 287 (47.6%) | 81 (22.2%) | 16 (15.8%) | <.001 |
| Race |  |  |  |  |  | <.001 |
| White | 113 (87.6%) | 172 (78.%) | 508 (84.5%) | 337 (92.3%) | 93 (93.0%) |  |
| Black/African American | 10 (7.8%) | 32 (14.7%) | 61 (10.1%) | 15 (4.1%) | 4 (4.0%) |  |
| Asian/Asian American | 4 (3.1%) | 8 (3.7%) | 12 (2.0%) | 4 (1.1%) | 1 (1.0%) |  |
| Other | 2 (1.6%) | 6 (2.8%) | 20 (3.3%) | 9 (2.5%) | 2 (2.0%) |  |
| Native lung disease category |  |  |  |  |  | <.001 |
| Obstructive/COPD | 39 (30.2%) | 89 (41.0%) | 157 (26.3%) | 101 (28.0%) | 36 (36.0%) |  |
| Pulmonary vascular disease/IPAH | 8 (6.2%) | 9 (4.1%) | 23 (3.9%) | 14 (3.9%) | 2 (2.0%) |  |
| Cystic fibrosis | 33 (25.6%) | 15 (6.9%) | 47 (7.9%) | 57 (15.8%) | 29 (29.0%) |  |
| Restrictive/IPF | 30 (23.3%) | 58 (26.7%) | 261 (43.7%) | 134 (37.1%) | 23 (23.0%) |  |
| Other | 19 (14.7%) | 46 (21.2%) | 109 (18.3%) | 55 (15.2%) | 10 (10.0%) |  |
| Transplant type |  |  |  |  |  | <.001 |
| Bilateral | 119 (93.0%) | 109 (50.5%) | 395 (66.2%) | 288 (80.0%) | 93 (93.0%) |  |
| Single | 9 (7.0%) | 107 (49.5%) | 202 (33.8%) | 72 (20.0%) | 7 (7.0%) |  |
| BMI, median (Q1, Q3), kg/m<sup>2</sup> | 23.5 (20.4, 27.1) | 25.9 (21.7, 29.8) | 26.0 (22.7, 28.9) | 25.3 (21.9, 28.7) | 23.9 (20.5, 27.0) | <.001 |
| Primary graft dysfunction grade 3 at 48 or 72 hours | 23 (18.4%) | 55 (26.7%) | 127 (22.2%) | 43 (12.6%) | 12 (12.4%) | <.001 |
| Baseline FEV1, median (Q1, Q3), L<sup>‡</sup> | 2.91 (2.38, 3.57) | 1.54 (1.32, 1.84) | 2.19 (1.91, 2.45) | 3.03 (2.80, 3.39) | 3.70 (2.82, 4.22) | <.001 |
| Baseline FVC, median (Q1, Q3), L* | 3.52 (2.85, 4.10) | 2.03 (1.78, 2.49) | 2.70 (2.38, 3.06) | 3.64 (3.26, 4.07) | 4.31 (3.32, 4.98) | <.001 |
| Baseline FEV1, median (Q1, Q3), % predicted<sup>‡</sup> | 87.7 (75.6, 104.1) | 60.0 (51.3, 73.3) | 75.1 (65.7, 88.0) | 93.5 (83.0, 108.0) | 100.3 (81.1, 115.4) | <.001 |
| Baseline FVC, median (Q1, Q3), % predicted<sup>‡</sup> | 84.2 (75.1, 95.3) | 65.4 (54.3, 76.4) | 74.1 (64.0, 86.7) | 87.9 (77.4, 99.3) | 91.1 (76.8, 104.6) | <.001 |
| **At CLAD onset or after CLAD** |  |  |  |  |  |  |
| Age at CLAD onset, median (Q1, Q3), yrs | 57.0 (35.0, 64.0) | 62.5 (53.5, 67.0) | 64.0 (56.0, 68.0) | 63.0 (51.0, 68.0) | 58.0 (40.0, 64.0) | <.001 |
| Transplant to CLAD, median (Q1, Q3), mos | 24.7 (13.9, 45.1) | 22.5 (12.9, 47.5) | 30.0 (13.7, 56.5) | 41.8 (20.7, 69.5) | 19.8 (6.4, 61.2) | <.001 |
| Early onset CLAD | 61 (47.3%) | 115 (52.3%) | 253 (42.0%) | 103 (28.2%) | 59 (58.4%) | <.001 |
| CLAD stage at onset |  |  |  |  |  | <.001 |
| Stage 1 | 99 (76.7%) | 125 (56.8%) | 504 (83.6%) | 336 (92.1%) | 81 (80.2%) |  |
| Stage 2 | 25 (19.4%) | 43 (19.5%) | 82 (13.6%) | 27 (7.4%) | 17 (16.8%) |  |
| Stage 3 | 5 (3.9%) | 37 (16.8%) | 16 (2.7%) | 2 (0.5%) | 2 (2.0%) |  |
| Stage 4 | 0 (0.0%) | 15 (6.8%) | 1 (0.2%) | 0 (0.0%) | 1 (1.0%) |  |
| FVC loss at CLAD onset | 46 (35.7%) | 124 (56.4%) | 213 (35.3%) | 103 (28.2%) | 22 (21.8%) | <.001 |
| FEV1/FVC ratio at CLAD onset, median (Q1, Q3) | 0.71 (0.62, 0.81) | 0.64 (0.50, 0.73) | 0.71 (0.62, 0.79) | 0.75 (0.67, 0.82) | 0.72 (0.62, 0.78) | <.001 |
| FEV1/FVC ratio at CLAD onset <0.7 | 62 (48.1%) | 146 (66.4%) | 288 (47.8%) | 120 (32.9%) | 44 (43.6%) | <.001 |
| FEV1 at CLAD onset, median (Q1, Q3), L | 2.06 (1.63, 2.41) | 0.99 (0.85, 1.13) | 1.57 (1.38, 1.77) | 2.27 (2.09, 2.53) | 2.65 (1.91, 3.18) | <.001 |
| FVC at CLAD onset, median (Q1, Q3), L | 2.84 (2.39, 3.52) | 1.57 (1.30, 1.92) | 2.22 (1.90, 2.63) | 3.13 (2.72, 3.57) | 3.73 (2.83, 4.53) | <.001 |
| Death or retransplantation within 9 months | 70 (54.3%) | 68 (30.9%) | 30 (5.0%) | 8 (2.2%) | 0 (0.0%) | <.001 |
| Mean of posterior probabilities | 0.84 | 0.77 | 0.77 | 0.77 | 0.82 |  |

\*Posttransplant baseline FVC is defined as the average of the two FVCs that pair with the two best posttransplant FEV1s measured at least three weeks apart
<sup>†</sup>% predicted values calculated using global lung function initiative (GLI) equation.<sup>24</sup>
<sup>‡</sup>Posttransplant baseline FEV1 is defined as the average of the two best posttransplant FEV1s measured at least three weeks apart.
Data presented as no. (%), unless otherwise indicated.
BMI indicates body mass index; CLAD, chronic lung allograft dysfunction; COPD, chronic obstructive pulmonary disease; FEV1, forced expiratory volume in 1 second; FVC, forced vital capacity;
IPAH, idiopathic pulmonary arterial hypertension; IPF, idiopathic pulmonary fibrosis.

#### Identification of early post-CLAD FEV1 trajectories in bilateral lung recipients

Interpreting pulmonary function in single lung recipients is complicated by the native lung, prompting some CLAD trials to focus on bilateral recipients. Therefore, we used data from 1021 bilateral recipients with probable CLAD in the LTOG cohort for latent class modeling in this subgroup (**Table E6**). Model selection resulted in a four-class solution based on fit statistics (**Tables E7 and E8**). The mean predicted post-CLAD FEV1 trajectories for each class are depicted in **Figure 3A**, while individual FEV1 trajectories using observed data are shown in **Figure E5**. **Figure 3B** illustrates cumulative incidence of graft loss. Notable clinical and physiological differences were found between the early post-CLAD FEV1 trajectory classes (**Table 4**). Classes one and two demonstrated poorer prognosis. Specifically, class one had the earliest onset of CLAD (median 22·6 months), advanced CLAD stages at presentation, and a predominantly restrictive ventilatory deficit (FVC loss at CLAD onset in 68·2%). All patients in class one experienced graft loss within nine months. Class two recipients were generally younger, enriched for cystic fibrosis, often displayed obstructive physiology at onset, and had an early post-CLAD graft loss rate of 16·7%. Conversely, class three was associated with a more favorable prognosis, a later onset of CLAD (median 38·2 months), a predominance of stage one disease at presentation (87·2%), higher baseline lung function, and superior lung function at CLAD onset. No class three patients experienced graft loss within nine months. Class four included individuals with the lowest baseline lung function, a high rate of PGD grade three (25·7%), and a reasonable short-term prognosis, with only 6·2% experiencing graft loss within nine months post-CLAD. These data demonstrate a clear risk gradient across lung function trajectory phenotypic classes in bilateral lung recipients.

**Figure 3.**
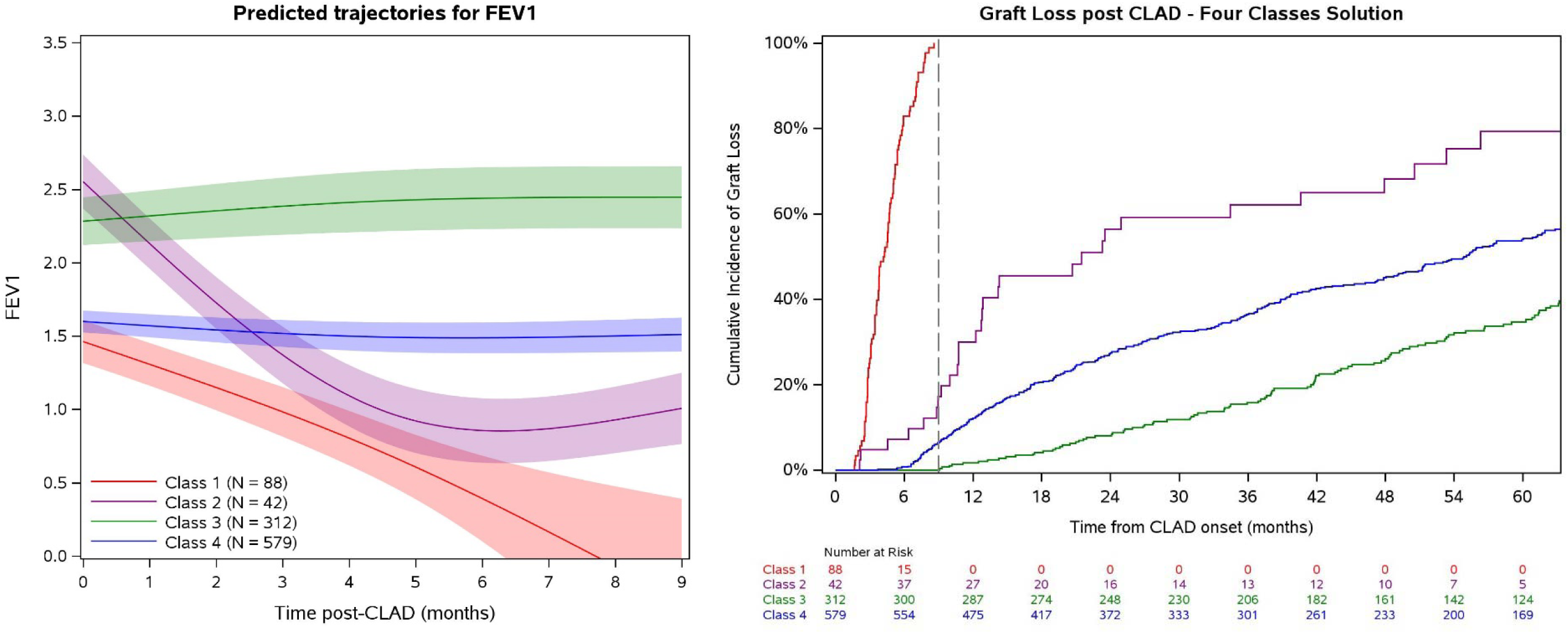
In the bilateral lung recipient LTOG CLAD cohort, **A)** estimated mean FEV1 trajectory in the nine months following CLAD onset for each of the four latent classes and **B)** cumulative incidence of graft loss after CLAD by class membership. Dashed line represents nine months post-CLAD.

**Table 4.**
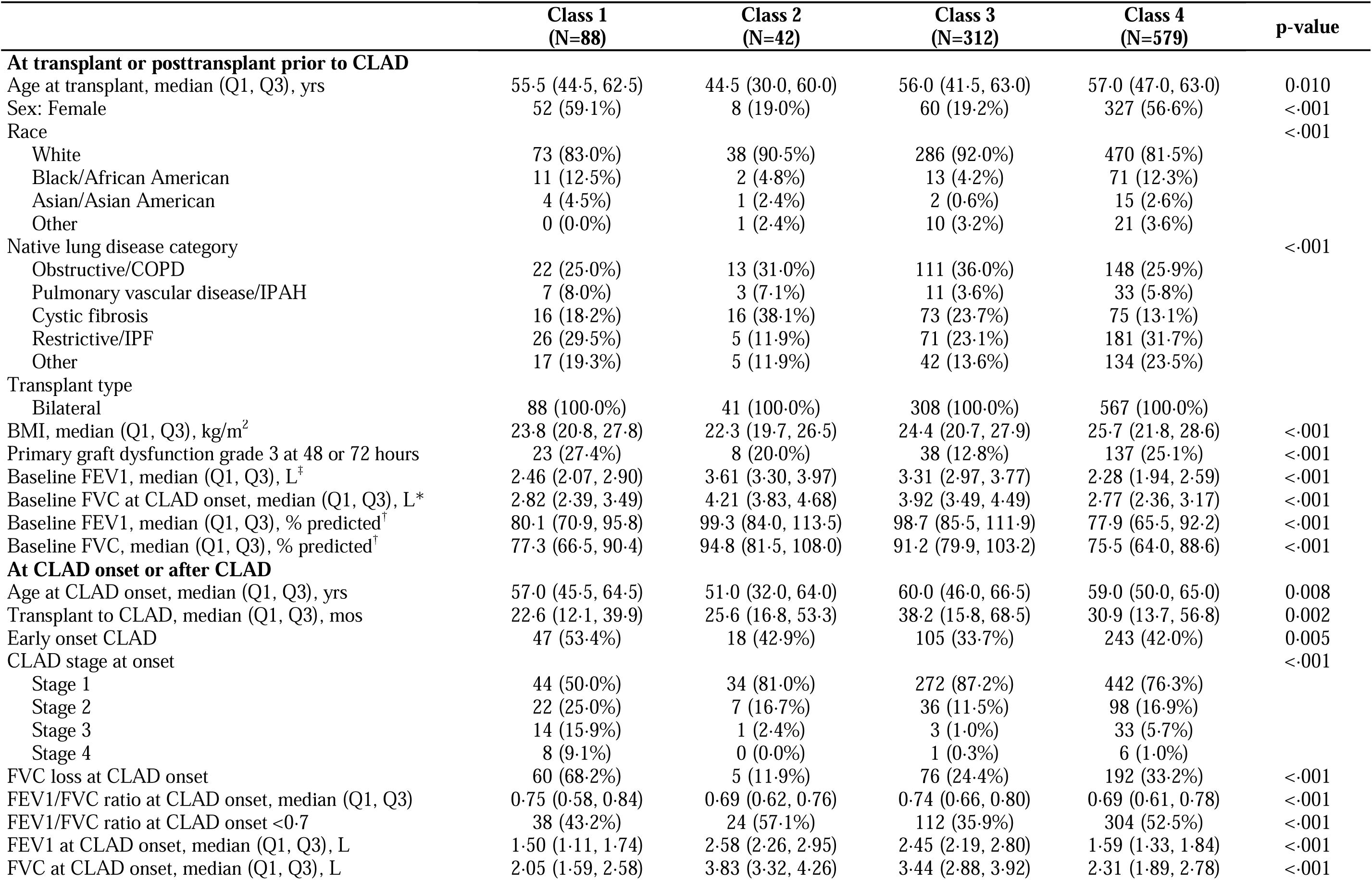
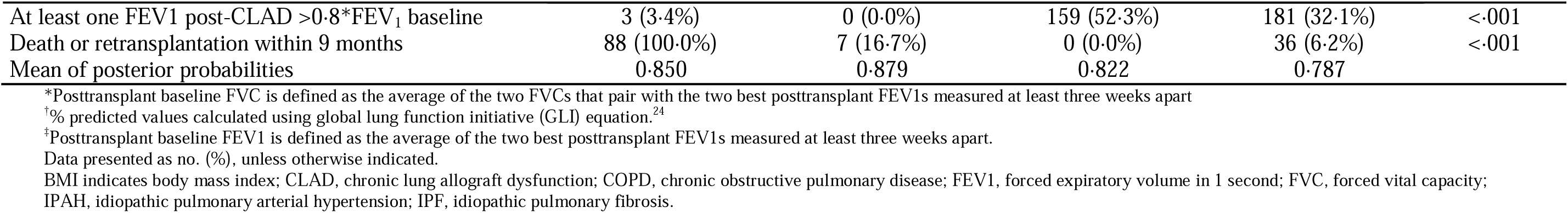
Clinical characteristics and CLAD onset features by class membership in the bilateral lung recipient LTOG probable CLAD cohort.

|  | **Class 1**<br>**(N=88)** | **Class 2**<br>**(N=42)** | **Class 3**<br>**(N=312)** | **Class 4**<br>**(N=579)** | **p-value** |
| --- | --- | --- | --- | --- | --- |
| **At transplant or posttransplant prior to CLAD** |  |  |  |  |  |
| Age at transplant, median (Q1, Q3), yrs | 55.5 (44.5, 62.5) | 44.5 (30.0, 60.0) | 56.0 (41.5, 63.0) | 57.0 (47.0, 63.0) | 0.010 |
| Sex: Female | 52 (59.1%) | 8 (19.0%) | 60 (19.2%) | 327 (56.6%) | <.001 |
| Race |  |  |  |  | <.001 |
| White | 73 (83.0%) | 38 (90.5%) | 286 (92.0%) | 470 (81.5%) |  |
| Black/African American | 11 (12.5%) | 2 (4.8%) | 13 (4.2%) | 71 (12.3%) |  |
| Asian/Asian American | 4 (4.5%) | 1 (2.4%) | 2 (0.6%) | 15 (2.6%) |  |
| Other | 0 (0.0%) | 1 (2.4%) | 10 (3.2%) | 21 (3.6%) |  |
| Native lung disease category |  |  |  |  | <.001 |
| Obstructive/COPD | 22 (25.0%) | 13 (31.0%) | 111 (36.0%) | 148 (25.9%) |  |
| Pulmonary vascular disease/IPAH | 7 (8.0%) | 3 (7.1%) | 11 (3.6%) | 33 (5.8%) |  |
| Cystic fibrosis | 16 (18.2%) | 16 (38.1%) | 73 (23.7%) | 75 (13.1%) |  |
| Restrictive/IPF | 26 (29.5%) | 5 (11.9%) | 71 (23.1%) | 181 (31.7%) |  |
| Other | 17 (19.3%) | 5 (11.9%) | 42 (13.6%) | 134 (23.5%) |  |
| Transplant type |  |  |  |  |  |
| Bilateral | 88 (100.0%) | 41 (100.0%) | 308 (100.0%) | 567 (100.0%) |  |
| BMI, median (Q1, Q3), kg/m<sup>2</sup> | 23.8 (20.8, 27.8) | 22.3 (19.7, 26.5) | 24.4 (20.7, 27.9) | 25.7 (21.8, 28.6) | <.001 |
| Primary graft dysfunction grade 3 at 48 or 72 hours | 23 (27.4%) | 8 (20.0%) | 38 (12.8%) | 137 (25.1%) | <.001 |
| Baseline FEV1, median (Q1, Q3), L<sup>†</sup> | 2.46 (2.07, 2.90) | 3.61 (3.30, 3.97) | 3.31 (2.97, 3.77) | 2.28 (1.94, 2.59) | <.001 |
| Baseline FVC at CLAD onset, median (Q1, Q3), L* | 2.82 (2.39, 3.49) | 4.21 (3.83, 4.68) | 3.92 (3.49, 4.49) | 2.77 (2.36, 3.17) | <.001 |
| Baseline FEV1, median (Q1, Q3), % predicted<sup>†</sup> | 80.1 (70.9, 95.8) | 99.3 (84.0, 113.5) | 98.7 (85.5, 111.9) | 77.9 (65.5, 92.2) | <.001 |
| Baseline FVC, median (Q1, Q3), % predicted<sup>†</sup> | 77.3 (66.5, 90.4) | 94.8 (81.5, 108.0) | 91.2 (79.9, 103.2) | 75.5 (64.0, 88.6) | <.001 |
| **At CLAD onset or after CLAD** |  |  |  |  |  |
| Age at CLAD onset, median (Q1, Q3), yrs | 57.0 (45.5, 64.5) | 51.0 (32.0, 64.0) | 60.0 (46.0, 66.5) | 59.0 (50.0, 65.0) | 0.008 |
| Transplant to CLAD, median (Q1, Q3), mos | 22.6 (12.1, 39.9) | 25.6 (16.8, 53.3) | 38.2 (15.8, 68.5) | 30.9 (13.7, 56.8) | 0.002 |
| Early onset CLAD | 47 (53.4%) | 18 (42.9%) | 105 (33.7%) | 243 (42.0%) | 0.005 |
| CLAD stage at onset |  |  |  |  | <.001 |
| Stage 1 | 44 (50.0%) | 34 (81.0%) | 272 (87.2%) | 442 (76.3%) |  |
| Stage 2 | 22 (25.0%) | 7 (16.7%) | 36 (11.5%) | 98 (16.9%) |  |
| Stage 3 | 14 (15.9%) | 1 (2.4%) | 3 (1.0%) | 33 (5.7%) |  |
| Stage 4 | 8 (9.1%) | 0 (0.0%) | 1 (0.3%) | 6 (1.0%) |  |
| FVC loss at CLAD onset | 60 (68.2%) | 5 (11.9%) | 76 (24.4%) | 192 (33.2%) | <.001 |
| FEV1/FVC ratio at CLAD onset, median (Q1, Q3) | 0.75 (0.58, 0.84) | 0.69 (0.62, 0.76) | 0.74 (0.66, 0.80) | 0.69 (0.61, 0.78) | <.001 |
| FEV1/FVC ratio at CLAD onset <0.7 | 38 (43.2%) | 24 (57.1%) | 112 (35.9%) | 304 (52.5%) | <.001 |
| FEV1 at CLAD onset, median (Q1, Q3), L | 1.50 (1.11, 1.74) | 2.58 (2.26, 2.95) | 2.45 (2.19, 2.80) | 1.59 (1.33, 1.84) | <.001 |
| FVC at CLAD onset, median (Q1, Q3), L | 2.05 (1.59, 2.58) | 3.83 (3.32, 4.26) | 3.44 (2.88, 3.92) | 2.31 (1.89, 2.78) | <.001 |
| At least one FEV1 post-CLAD >0.8*FEV<sub>1</sub> baseline | 3 (3.4%) | 0 (0.0%) | 159 (52.3%) | 181 (32.1%) | <.001 |
| Death or retransplantation within 9 months | 88 (100.0%) | 7 (16.7%) | 0 (0.0%) | 36 (6.2%) | <.001 |
| Mean of posterior probabilities | 0.850 | 0.879 | 0.822 | 0.787 |  |

\*Posttransplant baseline FVC is defined as the average of the two FVCs that pair with the two best posttransplant FEV1s measured at least three weeks apart
†% predicted values calculated using global lung function initiative (GLI) equation.<sup>24</sup>
‡Posttransplant baseline FEV1 is defined as the average of the two best posttransplant FEV1s measured at least three weeks apart.
Data presented as no. (%), unless otherwise indicated.
BMI indicates body mass index; CLAD, chronic lung allograft dysfunction; COPD, chronic obstructive pulmonary disease; FEV1, forced expiratory volume in 1 second; FVC, forced vital capacity;
IPAH, idiopathic pulmonary arterial hypertension; IPF, idiopathic pulmonary fibrosis.

### Classification of early post-CLAD FEV1 trajectories in bilateral lung recipients

We next used CART modeling to generate a clinically interpretable decision rule that could be used at CLAD onset or shortly thereafter to predict the early post-CLAD FEV1 progression course a patient may experience. The model was derived in the LTOG bilateral recipient CLAD cohort and then applied to the CTOT-20 bilateral recipient CLAD cohort (n=144) for validation. The performance characteristics of the clinical only and clinical+spirometry models in the derivation cohort are shown in **Table 5**. The model including the spirometric variables had superior discriminatory ability compared with the clinical-only model (AUC 0·85 vs. 0·68). The decision tree for the clinical+spirometry model is shown in **Figure 4**. The most informative features of the model were the values of the FEV1 at CLAD onset, the FEV1 that confirmed CLAD, and the posttransplant baseline FEV1 (**Table E9**). The clinical+spirometry model correctly classified a higher proportion of patients in the better prognosis groups (classes three and four: 88·5% and 91·9%) as compared with the poor prognosis groups (classes one and two: 31·8% and 47·6%).

**Table 5.** Performance characteristics of clinical only and clinical + PFT CART models in the derivation cohort.

|  | **AUC** | **Accuracy** | **Sensitivity** | **Specificity** |
| --- | --- | --- | --- | --- |
| Clinical only | 0.68 | 0.62 | 0.33 | 0.81 |
| Clinical + spirometry | 0.85 | 0.81 | 0.60 | 0.91 |

AUC indicates area under the receiver operating characteristic curve.

**Figure 4.**
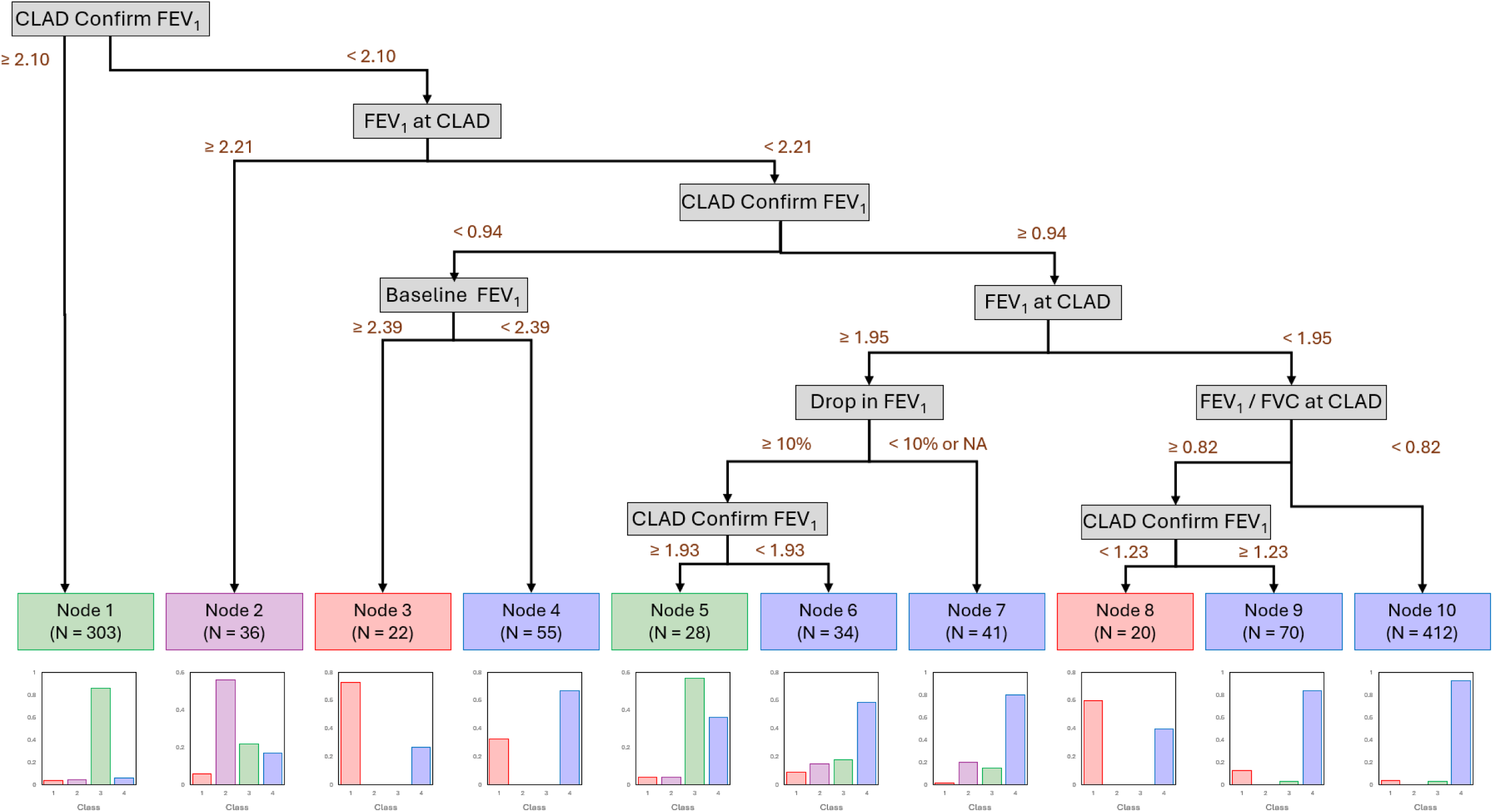
Decision tree for classification of bilateral lung recipients with probable CLAD to FEV1 trajectory phenotypes discovered in the latent class model for LTOG bilateral recipients with CLAD. Results of the clinical + spirometry model are shown. • CLAD Confirm FEV1 = the value of the FEV1 taken at least 21 days after the CLAD onset date that confirms probable CLAD state (in liters). • FEV1 at CLAD = the value of the FEV1 at CLAD onset (in liters). • Baseline FEV1 = the value of the baseline FEV1 (in liters), the baseline FEV1 is the average of the two highest FEV1s posttransplant taken at least 3 weeks apart. • Drop in FEV1 = the % decline from the last FEV1 prior to CLAD onset date to the CLAD onset FEV1 (no FEV1 in 6 months prior to CLAD = NA). • FEV1/FVC at CLAD = FEV1/FVC ratio at CLAD onset.

When the clinical+spirometry CART algorithm was applied to the CTOT-20 bilateral CLAD cohort, the predicted class distribution of the CTOT-20 patients was similar to both the observed and predicted class distribution in the LTOG bilateral recipients (**Table E10**). Moreover, when the FEV1 trajectories for the CTOT-20 bilateral CLAD patients were plotted based on the class assigned by the decision tree derived in LTOG bilateral CLAD patients, we observed a similar trajectory of phenotypes to those identified among the LTOG bilateral CLAD cohort, though confidence limits are wide which is reflective of the limited sample size (**Figure 5**). Together these data identify the most informative features of the FEV1 trajectory and support the robustness of the decision tree in distinguishing the early post-CLAD FEV1 trajectory.

**Figure 5.**
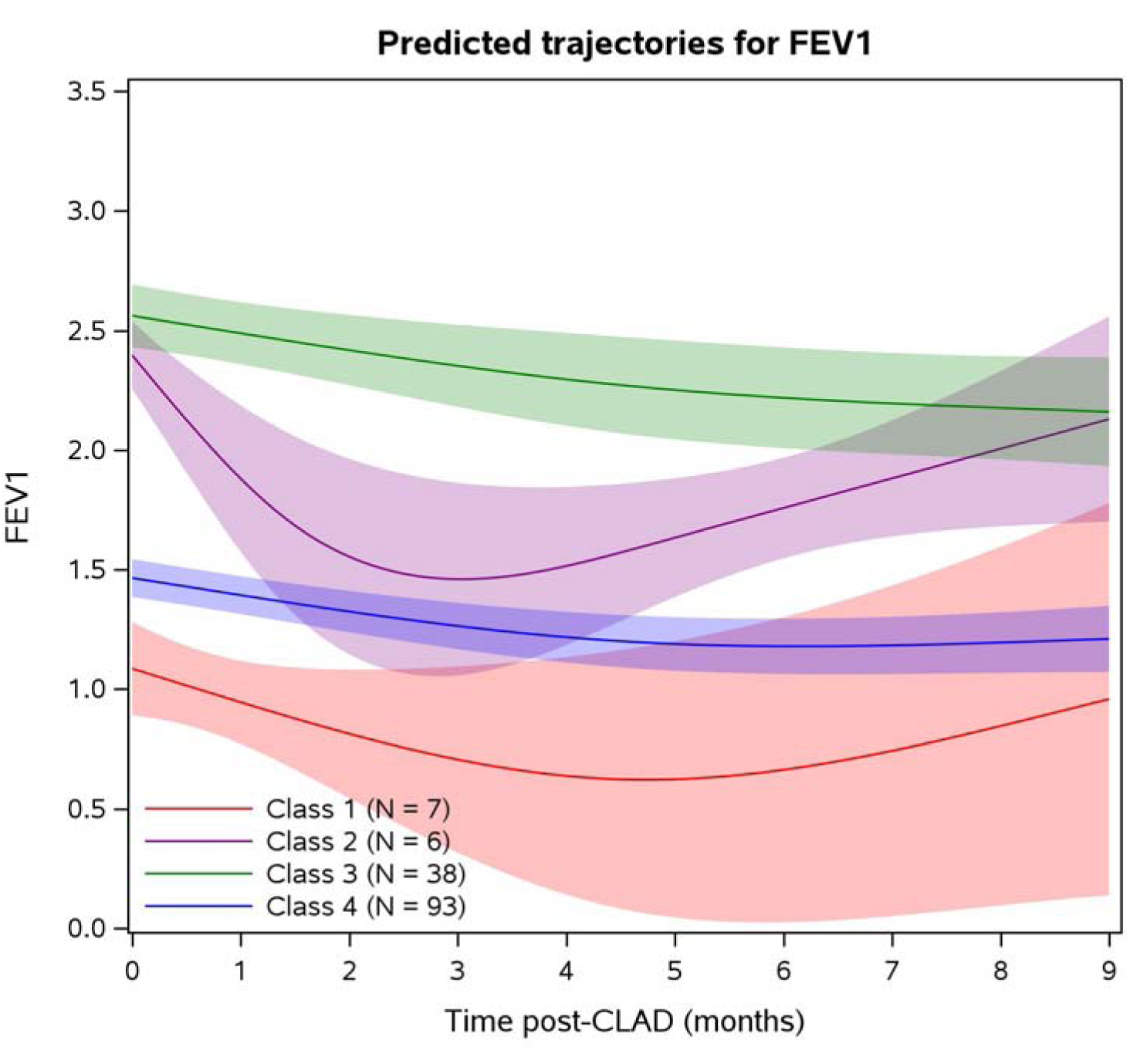
Estimated mean FEV1 trajectory for CTOT-20 bilateral lung recipients with CLAD according to the class assigned by the decision tree derived in the LTOG bilateral lung CLAD cohort.

## DISCUSSION

We found that early post-CLAD lung function behavior clusters into distinct, clinically meaningful FEV1 trajectory phenotypes. Across two largely independent prospective cohorts, latent class modeling identified reproducible subgroups spanning rapid early decline, intermediate patterns, and relatively stable courses; in the larger LTOG cohort, we additionally observed a subgroup with early improvement in FEV1 after probable CLAD onset. These trajectory classes were associated with a clear risk gradient for graft loss. Moreover, we generated a clinically interpretable classification rule in bilateral recipients that showed good class discrimination when spirometric features were included and also performed well in predicting FEV1 trajectory phenotypes in a new population. Collectively, these findings help resolve the heterogeneity in probable CLAD progression and suggest a method for predicting the lung function trajectory phenotype of prospective patients.

A central observation from our study is that the early trajectory after probable CLAD onset contains clinically actionable heterogeneity. We found a modest proportion of patients with CLAD (30·5% in CTOT-20, 24·6% in LTOG, and 12·7% among LTOG bilateral recipients) fell into the highest risk FEV1 trajectory phenotypes characterized by notable lung function decline and greater graft loss in the first nine months after CLAD. Female recipients were disproportionately represented in these higher risk trajectory classes. Additionally, sex was the most informative variable in discriminating the latent classes in the clinical-only CART model with higher variable importance than CLAD stage or timing of CLAD onset. These findings build on previous evidence of sex differences in CLAD; for example, our prior single-center data showed that patients with FVC loss at CLAD, which associates with poorer outcomes, are more likely to be female.^8^ This suggests biological factors like lung size and thus pulmonary reserve, immune response, or susceptibility to injury may impact early CLAD progression. We also found recipients with cystic fibrosis were consistently overrepresented in higher risk trajectory classes. Together these data suggest that females and recipients with cystic fibrosis meeting probable CLAD criteria in particular warrant close early clinical surveillance with more frequent spirometry and prioritization for early interventions.

The findings from our study extend established CLAD phenotyping approaches that distinguish obstructive and restrictive forms of disease. Prior work has shown that restrictive physiology and FVC loss at CLAD onset are associated with inferior outcomes compared with predominantly obstructive patterns.^6,8,9^ However, we found that presence of FVC loss and FEV1/FVC ratio <0·7 alone did not fully explain the wide variability in lung function trajectories observed after probable CLAD onset. This suggests that existing physiological phenotypic classifications, while clinically useful, are insufficient to capture the full spectrum of CLAD progression. Instead, our trajectory-based approach captures dynamic patterns of decline, stability, and improvement using routinely collected spirometry, offering a complementary lens through which to view early CLAD behavior. Such an approach may be particularly valuable for probable CLAD-focused interventional trials, where early enrollment is desired but population heterogeneity risks diluting treatment effects. Trajectory-informed stratification could improve trial efficiency while also enhancing patient prognostication.

In our pragmatic classification model, the most informative predictors were the FEV1 at CLAD onset, the FEV1 on the confirmatory spirometry, and the posttransplant baseline FEV1, indicating that the pulmonary reserve at posttransplant baseline and at CLAD onset in addition to the very near-term trajectory convey prognostic information. This finding reinforces the intent of the ISHLT diagnostic framework, in which persistence of decline increases diagnostic certainty^2^ and extends it by showing that the confirmatory PFT also helps define trajectory class. Notably the median time from the CLAD onset spirometry to the CLAD confirmatory spirometry was 42 days in both LTOG and CTOT-20 bilateral cohorts, reinforcing the need for early reassessment of spirometry after a threshold FEV1 decline has occurred. The importance of baseline lung function further suggests that pulmonary reserve influences the likelihood of CLAD progression, consistent with emerging data on baseline lung allograft dysfunction (BLAD). BLAD has been variably associated with subsequent CLAD development but has been more consistently linked to worse graft survival,^25–28^ and in our analysis patients with poorer baseline lung function were concentrated in CLAD FEV1 trajectory classes with less favorable prognosis.

While this study leverages large, prospective multicenter cohorts, some limitations warrant consideration. Spirometry was obtained as part of clinical care, introducing variability in testing frequency and timing. Probable CLAD ascertainment differed between cohorts, with prospective adjudication in CTOT-20 and algorithmic identification in LTOG, raising the possibility of residual misclassification despite the use of standardized spirometric criteria. Despite this, similar latent classes of FEV1 trajectories were identified across both datasets emphasizing the generalizability of these findings. We focused on the first nine months after CLAD onset to ensure sufficient data density and to minimize bias from early graft loss, but this limits inference about longer-term trajectories. Absolute FEV1 was utilized instead of percent predicted values, as the definition, staging, and management of CLAD rely on absolute measures. Similarly, most CLAD clinical trials have incorporated changes in absolute FEV1 as an inclusion for progressive CLAD. Consequently, absolute FEV1 constitutes a clinically relevant metric for monitoring disease progression. Models using percent predicted values may provide complementary information. Finally, while the CART model performed well overall, discrimination was strongest for more favorable prognosis classes, underscoring the need for additional external validation and potential incorporation of additional predictors.

In summary, we identified distinct and reproducible latent classes of early lung function progression following probable CLAD diagnosis. These trajectory-based phenotypes elucidate meaningful clinical heterogeneity beyond current clinical CLAD classifications, establish a framework for more accurate risk stratification, and may inform strategies for trial enrichment or stratification, as well as the development of individualized clinical monitoring plans. Notably, recipient factors, CLAD stage, and CLAD timing alone did not adequately distinguish the trajectory phenotypes; our findings instead underscore the significance of specific spirometric characteristics in determining patterns of early lung function progression after CLAD onset. Investigating the molecular and immunologic features associated with these divergent clinical trajectories may reveal novel therapeutic targets for CLAD and ultimately enhance lung transplant outcomes.

## Author contributions

All authors contributed to the design of the study; MLN, MRA, KA, JAB, LB MB, MC, GD, JYH, LK, TM, JM, MO, KP, JMR, JGR, DWR, PDS, JPS, LGS, LDS, WT, SSW, JDC, SMP, and JLT contributed to the acquisition of the data; MLN, DW, HH, PW, SMP and JLT contributed to the development of the analysis plan; and MLN and DW performed the data analysis. All authors contributed to the interpretation of the data. MLN and JLT drafted the manuscript. All authors had the opportunity to critically revise the manuscript and approve the submission.

## Data sharing statement

Deidentified analysis data may be made available upon reasonable request and written agreement by contacting the corresponding author.

## Declaration of interests

**Neely:** Support for the present manuscript from 5U01-HL145435, 5R21-AI168582, CF Foundation CTOT-20 Extension (Palmer 19AB0); Grants or contracts from 5U01-AI163099, 1U24-HL163122, Boehringer Ingelheim IPF/ILD-PRO Registries; Payment or honoraria from NC State University, *Journal of Heart and Lung Transplantation*; nothing else to disclose.

**Wojdyla:** To be collected

**Hong:** Support for the present manuscript from NIH R21AI168582; nothing else to disclose.

**Wang:** Nothing to disclose.

**Anderson:** To be collected.

**Arroyo:** Nothing to disclose.

**Belperio:** Nothing to disclose.

**Benvenuto:** To be collected.

**Budev:** To be collected.

**Combs:** To be collected.

**Dhillon:** Grants or contracts from NHLBI, CF Foundation; nothing else to disclose.

**Hsu:** Support for the present manuscript from U01-HL145435; Leadership or fiduciary role for National Kidney Foundation, Public Library of Science, American Medical Association; nothing else to disclose.

**Kalman:** Support for the present manuscript from NIH; nothing else to disclose.

**Martinu:** To be collected.

**McDyer:** To be collected.

**Oyster:** To be collected.

**Pandya:** To be collected.

**Reynolds:** To be collected.

**Rim:** Nothing to disclose.

**Roe:** Nothing to disclose.

**Shah:** To be collected.

**J. Singer:** Grants or contracts from U01-HL145435-06; nothing else to disclose.

**L. Singer:** Grants or contracts from Sanofi; Payment or honoraria from Sanofi; Participation on a data safety monitoring board or advisory board for Renovion, Zambon; nothing else to disclose.

**Snyder:** To be collected.

**Tsuang:** Nothing to disclose.

**Weigt:** Support for the present manuscript from National Institute of Allergy and Infectious Diseases (U01AI113315), Cystic Fibrosis Foundation (PALMER19AB0); Grants or contracts from CareDx, Zambon; Consulting fees from Boehringer Ingelheim Pharmaceuticals, Inc., Sanofi Pharmaceuticals; Payment or honoraria from Boehringer Ingelheim Pharmaceuticals, Inc., CareDx; Patents planned, issued or pending for U.S.Provisional Patent Application No. 63/862,482 entitled BIOMARKER FOR ALLOGRAFT INJURY OR REJECTION; nothing else to disclose.

**Christie:** Support for the present manuscript from NIH/NHLBI; Grants or contracts from NIH/NHLBI, CF Foundation; Consulting fees from GSK, United Health Care; Support for attending meetings and/or travel from International Society of Heart and Lung Transplantation; Participation on a data safety monitoring board or advisory board for NIH PETALnet, NHLBI; Other financial or non-financial interests include consulting as an expert witness in asbestos litigation for various law firms; nothing else to disclose.

**Palmer:** Grants or contracts from Sanofi, Veloxis, Bristol Myers Squibb, CareDx, Boehringer Ingelheim Pharmaceuticals, Inc.; Royalties or licenses from Up to Date; Consulting fees from Abbvie, Sanofi, Mallinckrodt Pharmaceuticals, Variant Bio, Incyte; Payment or honoraria from Sanofi, Boehringer Ingelheim Pharmaceuticals, Inc.; nothing else to disclose.

**Todd:** Support for the present manuscript from NIH/NHLBI, NIH/NIAID, Cystic Fibrosis Foundation; Grants or contracts from NIH, Boehringer Ingelheim, Cystic Fibrosis Foundation, Sanofi; Consulting fees from Sanofi; Participation on a data safety monitoring board or advisory board from Sanofi, Avalyn; nothing else to disclose.

