## Supplemental Appendix for "Latent Class Analysis Identifies Pulmonary Function Trajectory Phenotypes in Lung Transplant Recipients with Chronic Allograft Dysfunction"

**SUPPLEMENTARY MATERIAL**

### **Supplemental Methods**

1. ***Preliminary Analyses for Latent Class Model in CTOT-20 CLAD Cohort***

Preliminary analyses were performed to enable selection of correlation structure for the repeated FEV_1_ data (over and above the correlation captured by the subject-specific random effects included in the longitudinal model component), the functional form of time in the longitudinal model component and the shape of hazard function for the graft loss (survival) component of the joint model. We also evaluated the relationship between hazards functions for the different latent classes, and the effects of the variables on the survival process as these can be common across classes or class specific. The tables below show the results of these analyses.

**A. Selection of correlation structure**

| **Correlation structure** | **Number of parameters** | **log likelihood** |  | **BIC** |
| --- | --- | --- | --- | --- |
| Independence | 15 | -288·46 |  | 656·85 |
| Brownian | 16 | -285·49 |  | 656·23 |
| Autocorrelation | 17 | -285·43 |  | 661·44 |

**B. Selection of functional form of time in the longitudinal component**

| **Functional form** | **Number of parameters** | **log likelihood** | **BIC** |
| --- | --- | --- | --- |
| Restricted cubic spline | 15 | -288·46 | 656·85 |
| Linear | 11 | -345·64 | 749·88 |
| Quadratic | 15 | -288·13 | 656·19 |

**C. Selection of the shape of the hazard function**

| **Shape of hazard function** | **Number of parameters** | **log likelihood** | **BIC** |
| --- | --- | --- | --- |
| Weibull | 15 | -288·46 | 656·85 |
| Piecewise (quantiles) | 15 | -296·08 | 672·08 |
| Piecewise (equidistant) | 15 | -293·70 | 667·32 |
| Spline (manual knots) | 18 | -285·49 | 666·88 |
| Spline (equidistant knots) | 18 | -285·59 | 667·09 |

D. **Selection of the relationship between hazard functions in different classes**

| **Relationship between hazard functions across classes** | **Number of classes** | **Number of parameters** | **log likelihood** | **BIC** |
| --- | --- | --- | --- | --- |
| Proportional hazards | 2 | 21 | -261·33 | 634·55 |
| Class-specific hazards | 2 | 22 | -260·71 | 638·63 |
| Common hazard | 2 | 20 | -273·41 | 653·37 |

**E. Variables in the survival component (common across classes vs. class-specific)**

|  | **Number of classes** | **Number of parameters** | **log likelihood** | **BIC** |
| --- | --- | --- | --- | --- |
| Common across all classes | 2 | 21 | -261·33 | 634·55 |
| Age (class-specific) | 2 | 22 | -260·87 | 638·96 |
| Transplant timing (class-specific) | 2 | 22 | -261·33 | 639·87 |
| Transplant type (class-specific) | 2 | 22 | -261·29 | 639·79 |

Based on this information, we selected an independent structure for the correlation structure with a restricted cubic spline for the functional relationship and Weibull distribution for the hazard function. While Brownian correlation structure is numerically slightly better with respect to BIC, it includes one extra parameter which causes estimation problems in more complex models. Similarly, a quadratic functional form is also numerically better with respect to BIC, but a restricted cubic spline behaves better in the extreme of the time distribution. These data also suggest a proportional hazard model with common effect across classes for all the variables in the survival process part of the joint model.

2. ***Preliminary Analyses for Latent Class Model in LTOG CLAD cohort***

As for CTOT-20, similar preliminary analyses were performed in the LTOG CLAD cohort to guide construction of the latent class model in this dataset. The tables below show the results of these analyses for LTOG.

A. **Selection of correlation structure**

| **Correlation structure** | **Number of parameters** | **log likelihood** | **BIC** |
| --- | --- | --- | --- |
| Independence | 15 | -2465·8 | 5040·5 |
| Brownian | 16 | -2353·0 | 4822·0 |
| Autocorrelation | 17 | -2348·9 | 4821·2 |

B. **Selection of functional form of time in the longitudinal component**

| **Functional form*** | **Number of parameters** | **log likelihood** | **BIC** |
| --- | --- | --- | --- |
| Restricted cubic spline | 16 | -2353·0 | 4822·0 |
| Linear | 12 | -2452·1 | 4991·2 |
| Quadratic | 16 | -2358·5 | 4833·1 |

*All models with Brownian correlation structure

C. **Selection of the shape of the hazard function**

| **Shape of hazard function*** | **Number of parameters** | **log likelihood** | **BIC** |
| --- | --- | --- | --- |
| Weibull | 16 | -2353·0 | 4822·0 |
| Piecewise (quantiles) | 16 | -2373·0 | 4862·2 |
| Piecewise (equidistant) | 16 | -2365·6 | 4847·4 |
| Spline (manual knots) | 19 | -2349·0 | 4835·9 |
| Spline (equidistant knots) | 19 | -2348·5 | 4834·8 |

*All models with Brownian correlation structure

**D. Selection of the relationship between hazard functions in different classes**

| **Relationship between hazard functions across classes** | **Number of classes** | **Number of parameters** | **log likelihood** | **BIC** |
| --- | --- | --- | --- | --- |
| Proportional hazards | 2 | 22 | -2230·6 | 4620·9 |
| Class-specific hazards | 2 | 23 | -2230·5 | 4627·9 |
| Common hazard | 2 | 21 | -2264·2 | 4680·8 |

E. **Variables in the survival component (common across classes vs. class-specific)**

|  | **Number of classes** | **Number of parameters** | **log likelihood** | **BIC** |
| --- | --- | --- | --- | --- |
| Common across all classes | 2 | 22 | -2230·6 | 4620·9 |
| Age (class-specific) | 2 | 23 | -2228·2 | 4623·4 |
| Transplant timing (class-specific) | 2 | 23 | -2230·6 | 4628·1 |
| Transplant type (class-specific) | 2 | 23 | -2226·6 | 4620·0 |

Based on this information, for the LTOG latent class model we selected a Brownian motion correlation structure with a restricted cubic spline for the functional relationship and Weibull distribution for the hazard function. Comparing BICs, the autocorrelation structure is numerically slightly better but with one extra parameter which causes estimation problems in more complex models. This information also suggests a proportional hazard model with common effect across classes for all the variables in the survival component of the joint model.

3. ***Preliminary Analyses for Latent Class Model in Bilateral Lung Recipients in the LTOG CLAD Cohort***

As for CTOT-20 and LTOG CLAD cohorts, similar preliminary analyses were performed among the bilateral lung recipients in the LTOG CLAD cohort to guide construction of the latent class model in this clinically relevant subset. The tables below show the results of these analyses.

A. **Selection of correlation structure**

| **Correlation structure** | **Number of parameters** | **log likelihood** | **BIC** |
| --- | --- | --- | --- |
| Independence | 14 | -2090·0 | 4277·0 |
| Brownian | 15 | -1987·1 | 4078·1 |
| Autocorrelation | 16 | -5666·5 | 11443·9 |

B. **Selection of functional form of time in the longitudinal component**

| **Functional form*** | **Number of parameters** | **log likelihood** | **BIC** |
| --- | --- | --- | --- |
| Restricted cubic spline | 15 | -1987·1 | 4078·1 |
| Linear | 11 | -2069·7 | 4215·6 |
| Quadratic | 15 | -1990·9 | 4085·7 |

*All models with Brownian correlation structure

C. **Selection of the shape of the hazard function**

| **Shape of hazard function*** | **Number of parameters** | **log likelihood** | **BIC** |
| --- | --- | --- | --- |
| Weibull | 15 | -1987·1 | 4078·1 |
| Piecewise (quantiles) | 15 | -2003·7 | 4111·3 |
| Piecewise (equidistant) | 15 | -1997·4 | 4098·8 |
| Spline (manual knots) | 18 | -1981·4 | 4087·5 |
| Spline (equidistant knots) | 18 | -1980·8 | 4086·3 |

*All models with Brownian correlation structure

D. **Selection of the relationship between hazard functions in different classes**

| **Relationship between hazard functions across classes** | **Number of classes** | **Number of parameters** | **log likelihood** | **BIC** |
| --- | --- | --- | --- | --- |
| Proportional hazards | 2 | 21 | -1924·5 | 3994·5 |
| Class-specific hazards | 2 | 22 | -1906·1 | 3964·7 |
| Common hazard | 2 | 20 | -1936·8 | 4012·2 |

E. **Variables in the survival component (common across classes vs. class-specific)**

|  | **Number of classes** | **Number of parameters** | **log likelihood** | **BIC** |
| --- | --- | --- | --- | --- |
| Common across all classes | 2 | 21 | -1924·5 | 3994·5 |
| Age (class-specific) | 2 | Model did not converge | | |
| Transplant timing (class-specific) | 2 | 22 | -1908·1 | 3968·6 |

Based on tables above, a restricted cubic spline relationship is the best for the functional form. Class-specific hazards seem better than proportional hazards, but the models don't converge when the number of classes is three or more. Similarly, a class-specific transplant timing effect seems better than a common effect for the two classes solution, but the common effect is better for three or more classes solutions. All models will consider a Brownian motion correlation matrix, with a Weibull hazard and common variable effects across classes.

### **Table E1. Candidate Variables for Classification and Regression Tree (CART) Analysis**

| **Clinical Only** | **Clinical + Spirometry** |
| --- | --- |
| Age at transplant | All clinical variables from the clinical only model |
| Age at CLAD diagnosis | FEV_1_ at CLAD diagnosis (liters) |
| Sex | FVC at CLAD diagnosis (liters) |
| Race | Baseline FEV_1_ (liters) |
| Hispanic ethnicity | Baseline FVC (liters) |
| Native lung disease (restrictive, obstructive, other) | FVC loss at CLAD onset (yes, no) |
| Body mass index at transplant | FEV_1_/FVC ratio at CLAD onset (continuous) |
| Early onset CLAD (yes/no) | FEV_1_/FVC ratio at CLAD onset <0·7 vs. ≥0·7 |
| Time from transplant to CLAD onset | Change from last FEV_1_ prior to CLAD onset to CLAD onset FEV_1_ (≥20% decline, 10–20% decline, <10% decline or increase, no FEV_1_ in 6 months prior to CLAD) |
| CLAD stage at diagnosis (1, 2, 3/4) | CLAD confirming FEV_1_ (liters) |

CLAD indicates chronic lung allograft dysfunction; FEV_1_, forced expiratory volume in 1 second; FVC, forced vital capacity.

### **Table E2. Model fit statistics for selection of number of classes in CTOT-20**

| **Class(es)** | **Number of Parameters** | **Log Likelihood** | **BIC** | **Entropy** | **Percentage of Patients in Each Class** | | | |
| --- | --- | --- | --- | --- | --- | --- | --- | --- |
|  |  |  |  |  | **Class 1** | **Class 2** | **Class 3** | **Class 4** |
| **1** | 15 | -288·46 | 656·85 | 1·000 | 100·0 |  |  |  |
| **2** | 21 | -261·33 | 634·55 | 0·889 | 18·4 | 81·6 |  |  |
| **3** | 27 | -246·76 | 637·37 | 0·684 | 54·9 | 23·3 | 21·8 |  |
| **4** | 33 | -237·24 | 650·31 | 0·730 | 43·7 | 22·8 | 7·8 | 25·7 |

BIC indicates Bayesian Information Criterion.

### **Table E3. Estimated parameters for the longitudinal model in the four classes solution in the CTOT-20 CLAD cohort**

|  | **Estimate** | **SE** | **p-value** |
| --- | --- | --- | --- |
| Intercept Class 1 | 1·6513 | 0·0970 | < 0·0001 |
| Intercept Class 2 | 0·7720 | 0·0558 | < 0·0001 |
| Intercept Class 3 | 1·5352 | 0·0804 | < 0·0001 |
| Intercept Class 4 | 2·1797 | 0·1081 | < 0·0001 |
| RCS(time) 1 Class 1 | -0·1916 | 0·0263 | < 0·0001 |
| RCS(time) 1 Class 2 | -0·0694 | 0·0233 | 0·0029 |
| RCS(time) 1 Class 3 | -0·0255 | 0·0128 | 0·0473 |
| RCS(time) 1 Class 4 | -0·0475 | 0·0239 | 0·0464 |
| RCS(time) 2 Class 1 | 0·0962 | 0·0294 | 0·0011 |
| RCS(time) 2 Class 2 | 0·0866 | 0·0322 | 0·0071 |
| RCS(time) 2 Class 3 | 0·0083 | 0·0155 | 0·5905 |
| RCS(time) 2 Class 4 | 0·0631 | 0·0301 | 0·0364 |

RCS indicates restricted cubic spline; SE, standard error.

### **Table E4. Model fit statistics for selection of number of classes in LTOG**

| **Class(es)** | **Number of Parameters** | **Log Likelihood** | **BIC** | **Entropy** | **Percentage of Patients in Each Class** | | | | | |
| --- | --- | --- | --- | --- | --- | --- | --- | --- | --- | --- |
|  |  |  |  |  | **Class 1** | **Class 2** | **Class 3** | **Class 4** | **Class 5** | **Class 6** |
| **1** | 16 | -2355·1 | 4826·3 | 1·000 | 100·0 |  |  |  |  |  |
| **2** | 22 | -2230·6 | 4620·9 | 0·888 | 7·5 | 92·5 |  |  |  |  |
| **3** | 28 | -2151·4 | 4505·9 | 0·601 | 7·1 | 59·0 | 33·9 |  |  |  |
| **4** | 34 | -2106·3 | 4459·3 | 0·663 | 9·2 | 49·4 | 36·2 | 5·2 |  |  |
| **5** | 40 | -2077·5 | 4445·2 | 0·660 | 9·1 | 15·5 | 25·7 | 42·5 | 7·1 |  |
| **6** | 46 | -2059·2 | 4452·3 | 0·672 | 10·6 | 31·7 | 13·5 | 12·3 | 24·8 | 7·3 |

BIC indicates Bayesian Information Criterion.

### **Table E5. Estimated parameters for the longitudinal model in the five classes solution in the LTOG CLAD cohort**

|  | **Estimate** | **SE** | **p-value** |
| --- | --- | --- | --- |
| Intercept Class 1 | 1·9903 | 0·0771 | < 0·0001 |
| Intercept Class 2 | 1·0224 | 0·0529 | < 0·0001 |
| Intercept Class 3 | 1·5544 | 0·0545 | < 0·0001 |
| Intercept Class 4 | 2·2341 | 0·0702 | < 0·0001 |
| Intercept Class 5 | 2·4416 | 0·0950 | < 0·0001 |
| RCS(time) 1 Class 1 | -0·2658 | 0·0223 | < 0·0001 |
| RCS(time) 1 Class 2 | -0·0270 | 0·0111 | 0·0152 |
| RCS(time) 1 Class 3 | -0·0082 | 0·0060 | 0·1738 |
| RCS(time) 1 Class 4 | 0·0156 | 0·0087 | 0·0721 |
| RCS(time) 1 Class 5 | 0·0635 | 0·0228 | 0·0053 |
| RCS(time) 2 Class 1 | 0·1740 | 0·0218 | < 0·0001 |
| RCS(time) 2 Class 2 | 0·0230 | 0·0121 | 0·0577 |
| RCS(time) 2 Class 3 | 0·0045 | 0·0066 | 0·4991 |
| RCS(time) 2 Class 4 | -0·0146 | 0·0097 | 0·1317 |
| RCS(time) 2 Class 5 | 0·0051 | 0·0223 | 0·8203 |

RCS indicates restricted cubic spline; SE, standard error.

### **Table E6. Characteristics of the bilateral lung recipients in the LTOG CLAD cohort**

|  | **(N=1021)** |
| --- | --- |
| Age at transplant, median (Q1, Q3), yrs | 57·0 (44·0, 63·0) |
| Age at CLAD onset, median (Q1, Q3), yrs | 59·0 (47·0, 66·0) |
| Sex: Female | 447 (43·8%) |
| Race |  |
| White | 867 (85·2%) |
| Black/African American | 97 (9·5%) |
| Asian/Asian American | 22 (2·2%) |
| Other | 32 (3·1%) |
| Hispanic | 48 (4·7%) |
| Native lung disease category |  |
| Obstructive/COPD | 294 (29·1%) |
| Pulmonary vascular disease/IPAH | 54 (5·4%) |
| Cystic fibrosis | 180 (17·8%) |
| Restrictive/IPF | 283 (28·0%) |
| Other | 198 (19·6%) |
| Transplant type |  |
| Bilateral | 1004 (100·0%) |
| BMI, median (Q1, Q3), kg/m^2^ | 24·9 (21·3, 28·3) |
| Primary graft dysfunction grade 3 at 48 or 72 hours | 206 (21·3%) |
| Transplant to CLAD, median (Q1, Q3), mos | 31·7 (14·0, 58·0) |
| Early onset CLAD | 413 (40·5%) |
| CLAD stage at onset |  |
| Stage 1 | 792 (77·6%) |
| Stage 2 | 163 (16·0%) |
| Stage 3 | 51 (5·0%) |
| Stage 4 | 15 (1·5%) |
| FVC loss at CLAD onset | 333 (32·6%) |
| FEV_1_/FVC ratio at CLAD onset, median (Q1, Q3) | 0·71 (0·62, 0·80) |
| FEV_1_/FVC ratio at CLAD onset <0·7 | 478 (46·8%) |
| FEV_1_ at CLAD onset, median (Q1, Q3), L | 1·83 (1·45, 2·26) |
| FVC at CLAD onset, median (Q1, Q3), L | 2·63 (2·09, 3·26) |
| Baseline FEV_1_, median (Q1, Q3), L | 2·60 (2·16, 3·17) |
| Baseline FEV_1_, median (Q1, Q3), % predicted | 85·3 (71·6, 100·8) |
| Baseline FVC, median (Q1, Q3), L | 3·14 (2·59, 3·79) |
| Baseline FVC, median (Q1, Q3), % predicted | 81·3 (68·7, 94·6) |

Data presented as no. (%), unless otherwise indicated.

BMI indicates body mass index; CLAD, chronic lung allograft dysfunction; COPD, chronic obstructive pulmonary disease; FEV_1_, forced expiratory volume in 1 second; FVC, forced vital capacity; IPAH, idiopathic pulmonary arterial hypertension; IPF, idiopathic pulmonary fibrosis.

### **Table E7. Model fit statistics for bilateral lung recipients in LTOG CLAD cohort**

| **Class(es)** | **Number of Parameters** | **Log Likelihood** | **BIC** | **Entropy** | **Percentage of Patients in Each Class** | | | | |
| --- | --- | --- | --- | --- | --- | --- | --- | --- | --- |
|  |  |  |  |  | **Class 1** | **Class 2** | **Class 3** | **Class 4** | **Class 5** |
| **1** | 15 | -1987·1 | 4078·1 | 1·000 | 100·0 |  |  |  |  |
| **2** | 21 | -1924·5 | 3994·5 | 0·383 | 52·7 | 47·3 |  |  |  |
| **3** | 27 | -1844·6 | 3876·2 | 0·901 | 11·0 | 84·9 | 4·1 |  |  |
| **4** | 33 | -1807·3 | 3843·2 | 0·680 | 8·6 | 30·6 | 4·1 | 56·7 |  |
| **5** | 39 | -1792·2 | 3854·6 | 0·720 | 8·5 | 34·0 | 4·5 | 49·7 | 3·0 |

BIC indicates Bayesian Information Criterion.

### **Table E8. Estimated parameters for the longitudinal model in the four classes solution among the bilateral lung recipients in the LTOG CLAD cohort**

|  | **Estimate** | **SE** | **p-value** |
| --- | --- | --- | --- |
| Intercept Class 1 | 1·4624 | 0·0746 | < 0·0001 |
| Intercept Class 2 | 2·5523 | 0·0941 | < 0·0001 |
| Intercept Class 3 | 2·2823 | 0·0831 | < 0·0001 |
| Intercept Class 4 | 1·5991 | 0·0384 | < 0·0001 |
| RCS(time) 1 Class 1 | -0·1549 | 0·0220 | < 0·0001 |
| RCS(time) 1 Class 2 | -0·4228 | 0·0296 | < 0·0001 |
| RCS(time) 1 Class 3 | 0·0361 | 0·0117 | 0·0020 |
| RCS(time) 1 Class 4 | -0·0295 | 0·0082 | 0·0003 |
| RCS(time) 2 Class 1 | -0·0559 | 0·0624 | 0·3702 |
| RCS(time) 2 Class 2 | 0·3347 | 0·0333 | < 0·0001 |
| RCS(time) 2 Class 3 | -0·0239 | 0·0124 | 0·0530 |
| RCS(time) 2 Class 4 | 0·0262 | 0·0089 | 0·0032 |

RCS indicates restricted cubic spline; SE, standard error.

### **Table E9. Variable importance in CART analysis for LTOG bilateral lung recipient CLAD cohort**

| **Variable** | **Clinical Only** | **Clinical+Spirometry** |
| --- | --- | --- |
| BMI | 51·8 | 1·6 |
| Sex | 100·0 | 0·6 |
| CLAD stage at onset | 71·2 | 2·2 |
| Time from transplant to CLAD | 76·3 | 1·5 |
| Age at CLAD | 32·6 | 1·3 |
| Age at Transplant | 54·3 | 1·3 |
| Race | 42·8 | 0·0 |
| Hispanic | 0·0 | 0·0 |
| Native Lung Disease | 43·7 | 0·0 |
| Early onset CLAD vs· not | 0·0 | 0·0 |
| Change from last FEV_1_ prior to CLAD onset to CLAD onset FEV_1_ |  | 1·3 |
| FEV_1_ at CLAD onset (liters) |  | 96·1 |
| Baseline FEV_1_ (liters) |  | 67·6 |
| FEV_1_/FVC ratio at CLAD onset |  | 1·9 |
| FEV_1_/FVC ration at CLAD onset <0·7 vs. ≥0·7 |  | 0·0 |
| FVC at CLAD onset (liters) |  | 5·5 |
| Baseline FVC (liters) |  | 67·6 |
| FVC loss at CLAD onset, yes vs. no |  | 1·1 |
| CLAD confirming FEV_1_ (liters) |  | 100·0 |

BMI indicates body mass index; CLAD, chronic lung allograft dysfunction; FEV_1_, forced expiratory volume in one second; FVC, forced vital capacity; PFT, pulmonary function test.

### **Table E10. Distribution of observed and predicted classes for bilateral lung recipients with CLAD**

| **Class** | **Observed (Classified by Latent Class Analysis in LTOG)** | **Predicted by Clincal+Spirometry CART in LTOG** | **Predicted by**  **Clinical +Spirometry**  **CART in CTOT-20** |
| --- | --- | --- | --- |
| 1 (Red) | 88 (8·6%) | 42 (4·1%) | 7 (4·9%) |
| 2 (Purple) | 42 (4·1%) | 36 (3·5%) | 6 (4·2%) |
| 3 (Green) | 312 (30·6%) | 331 (32·4%) | 38 (26·4%) |
| 4 (Blue) | 579 (56·7%) | 612 (59·9%) | 93 (64·6%) |
| **Total** | 1021 (100·0%) | 1021 (100·0%) | 144 (100·0%) |

CART indicates classification and regression tree; CTOT-20, Clinical Trials in Organ Transplantation; LTOG, Lung Transplant Outcomes Group.

### **Figure E1. Predicted mean FEV_1_ trajectory in the nine months following CLAD onset for two class solution in CTOT-20 CLAD cohort**

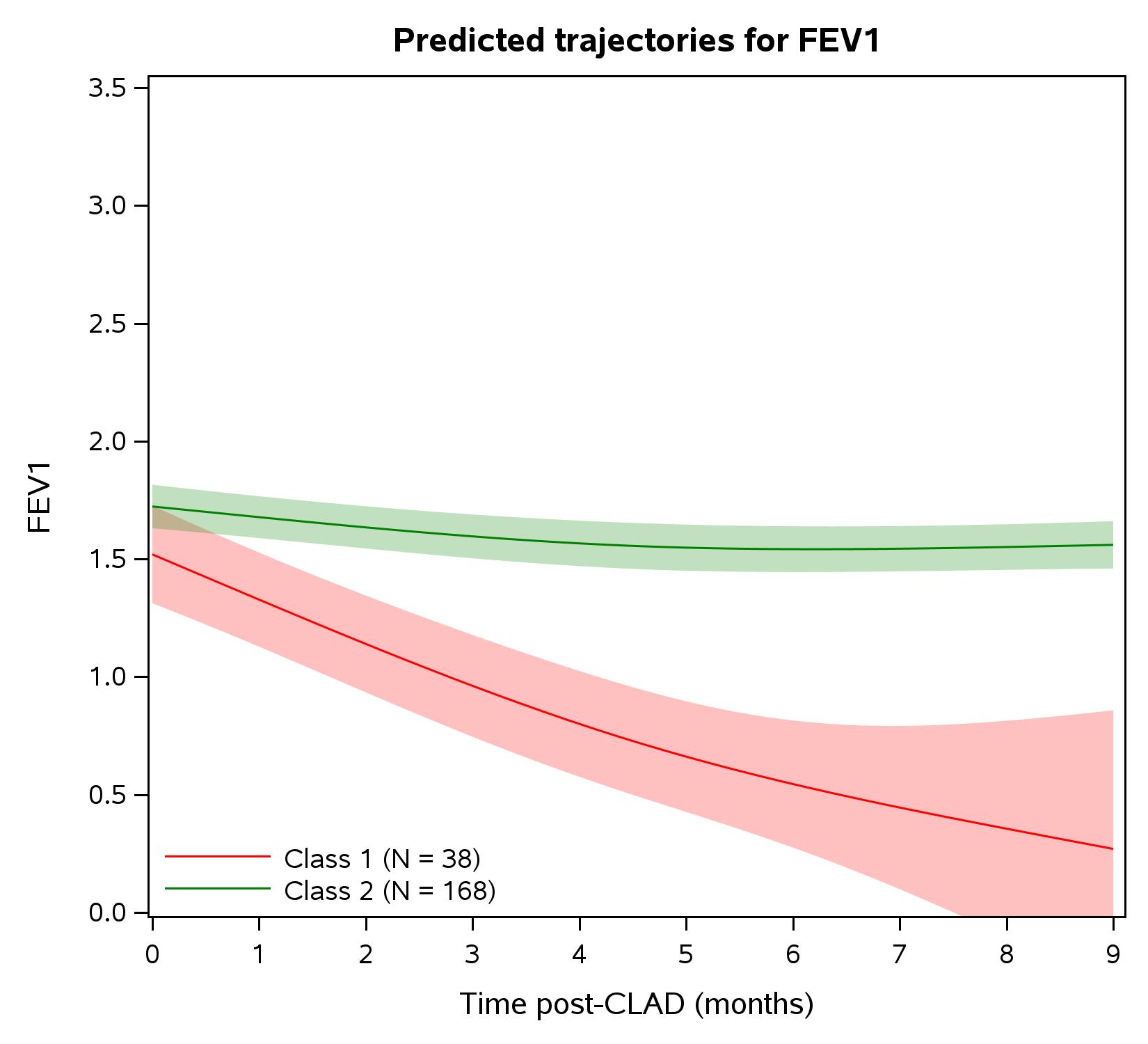

### **Figure E2. Predicted mean FEV_1_ trajectory in the nine months following CLAD onset for three class solution in CTOT-20 CLAD cohort**

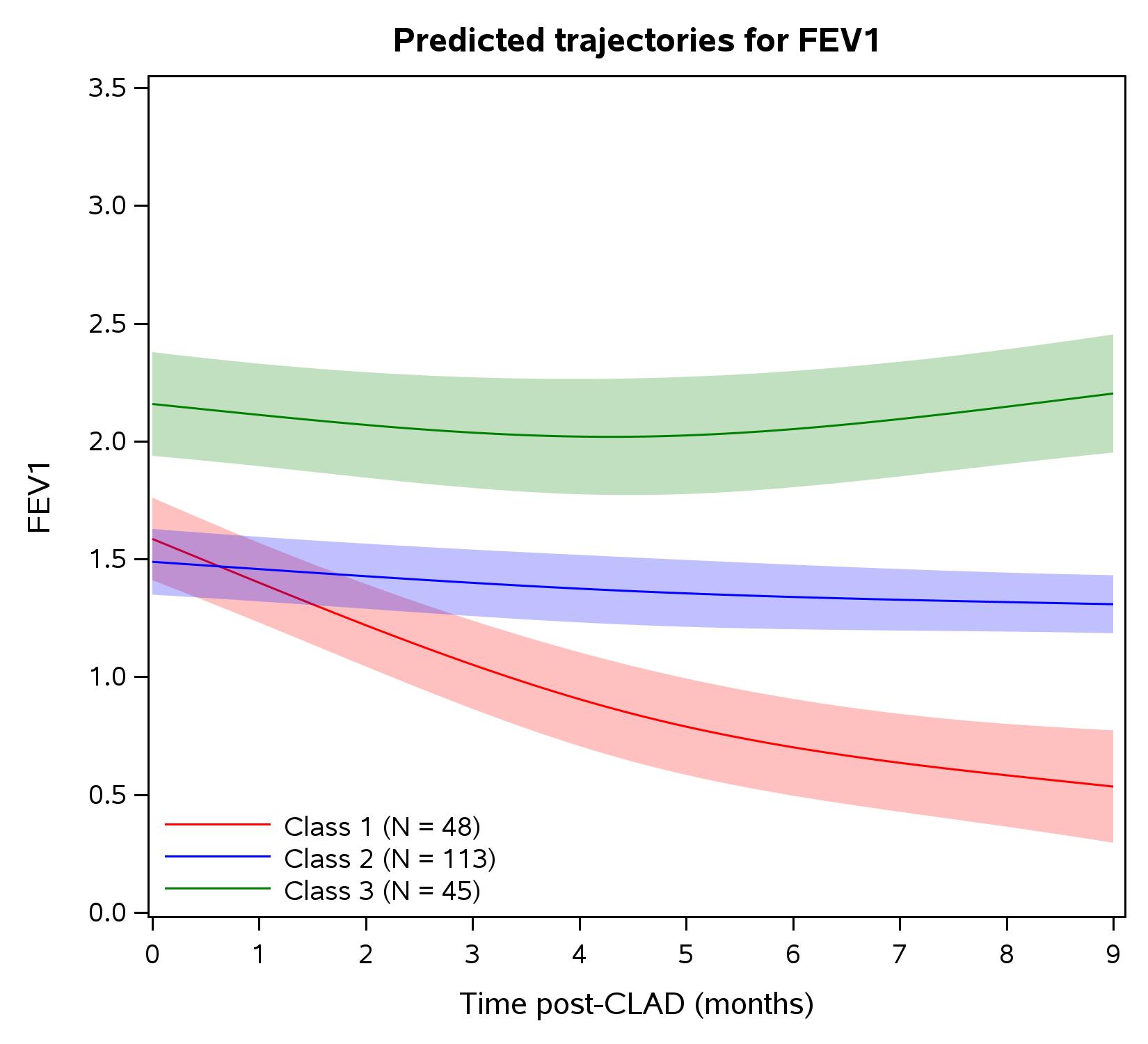

### **Figure E3. Observed individual FEV_1_ trajectories in the nine months following CLAD onset for the four classes solution in CTOT-20 CLAD cohort. The mean predicted FEV_1_ trajectory line for each class is overlaid**

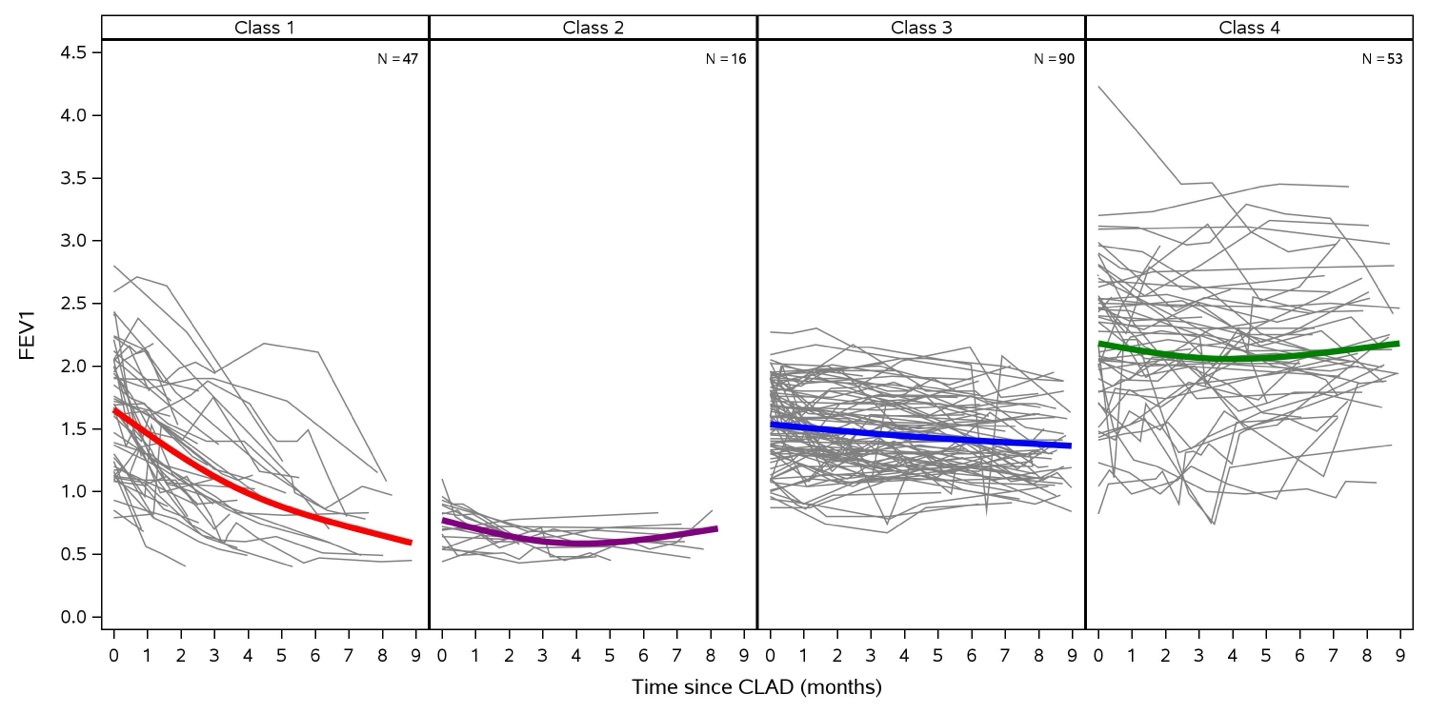

### **Figure E4. Observed individual FEV_1_ trajectories in the nine months following CLAD onset for the five classes solution in LTOG CLAD cohort. The mean predicted FEV_1_ trajectory line for each class is overlaid**

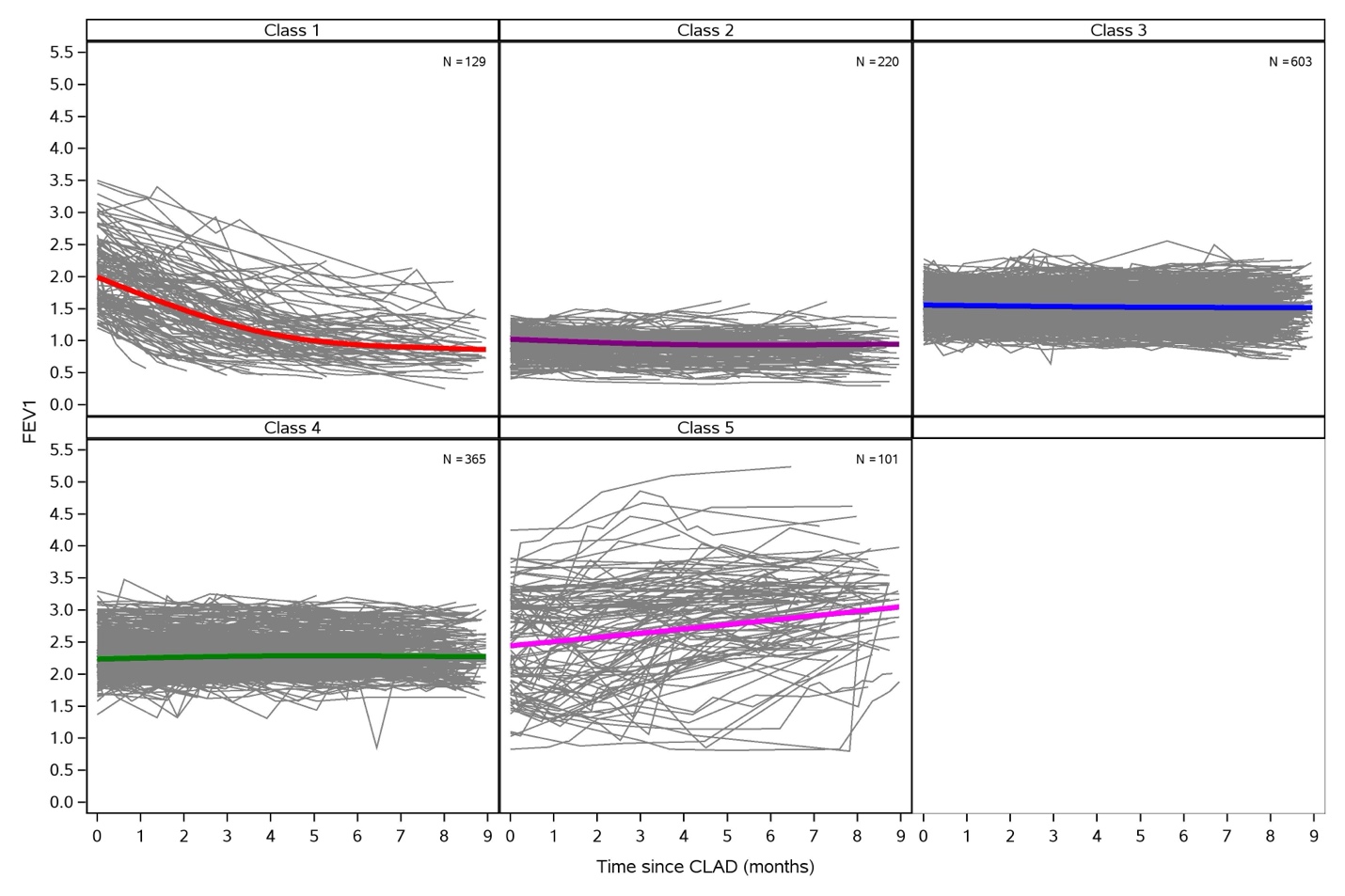

### **Figure E5. Observed individual FEV_1_ trajectories in the nine months following CLAD onset for the four classes solution among the bilateral lung recipients in the LTOG CLAD cohort. The mean predicted FEV_1_ trajectory line for each class is overlaid**

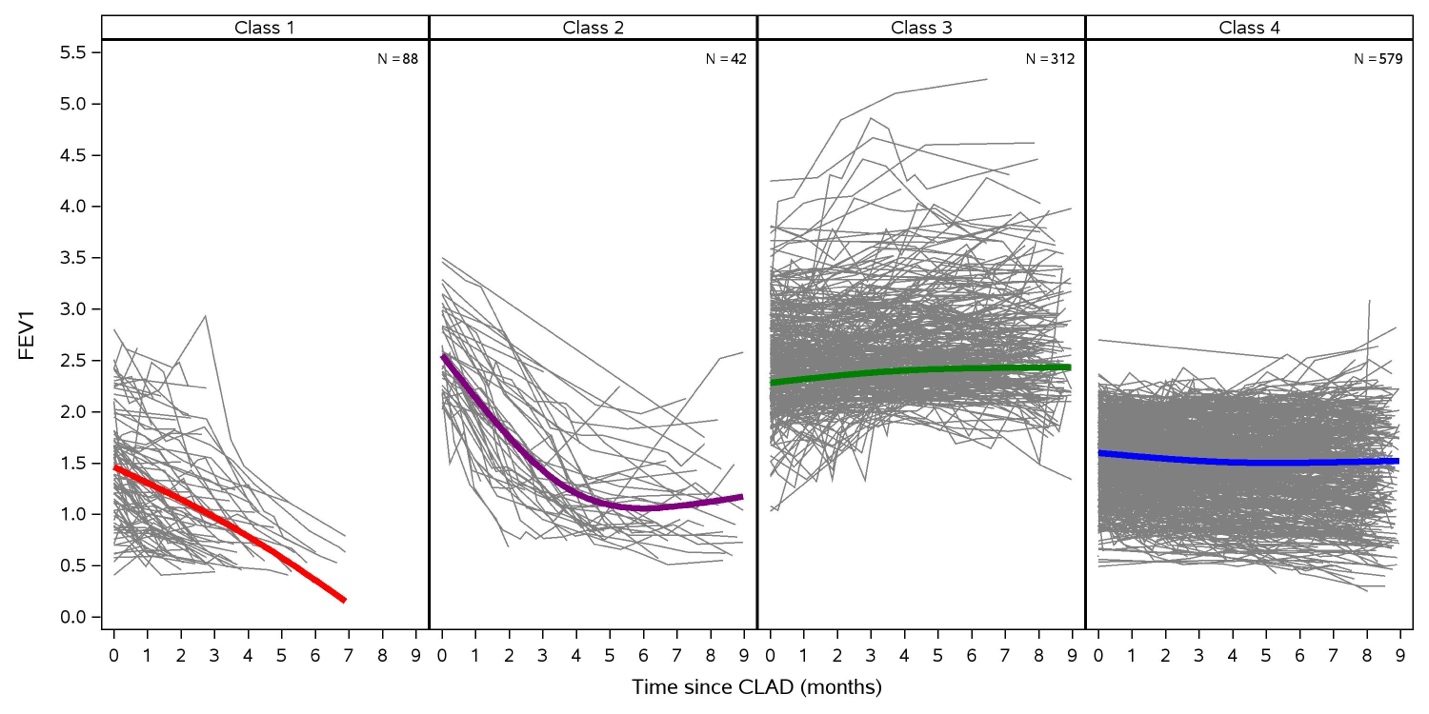
